# Exploring the genetic architecture of multimorbidity and its impact on long COVID risk

**DOI:** 10.64898/2026.05.18.26353402

**Authors:** Sasha Bauer, Ruth C. E. Bowyer, Laura Bravo Merodio, Georgios Gkoutos, Davide Vetrano, Thomas Jackson, Claire J. Steves, Maxim B. Freidin

## Abstract

Multimorbidity, the co-occurrence of multiple long-term conditions, represents a major challenge for ageing populations, yet its genetic architecture and relationship to long COVID remain unclear, despite shared epidemiological risk factors. We analysed multimorbidity patterns in 86,756 White British UK Biobank participants aged ≥65 years, identifying six clusters spanning neurodegenerative, cardiac, gastrointestinal, musculoskeletal, vascular, and cancer & eye disease domains. Genome-wide association studies and post-GWAS analyses revealed significant loci in five clusters, including *APOE, LPA*, and *CDKN2B-AS1*, with patterns of genetic correlation consistent with known disease relationships. Notably, a shared variant within the *APOE-APOC1* locus showed opposite effect directions for the musculoskeletal and vascular clusters, consistent with their negative genetic correlation. Investigating the multimorbidity-long COVID relationship via genetic correlation and Mendelian randomisation revealed no evidence of significant shared genetic architecture or causal effects. These findings indicate that multimorbidity clusters represent biologically structured, partly heritable phenotypes, whereas genetic overlap with long COVID appears limited.

## INTRODUCTION

Long COVID refers to persisting or newly developed symptoms following acute infection with SARS-CoV-2 which cannot be explained by an alternative diagnosis^1–4^. Definitions vary across health authorities: the UK National Institute for Health and Care Excellence (NICE) includes both “ongoing symptomatic COVID-19” (symptoms lasting 4 to 12 weeks) and “post-COVID-19 syndrome” (symptoms persisting beyond 12 weeks), whilst the World Health Organisation defines long COVID as symptoms lasting at least 8 weeks after infection^3–5^. Prevalence estimates vary widely, reflecting this lack of consensus regarding long COVID definition and methodological heterogeneity across studies. Using a 3-month threshold, the Global Burden of Disease study estimated that 6.2% (2.4%-13.3%) of individuals experienced persistent symptoms following COVID-19 infection globally in 2020-2021^6,7^. In the UK, as of March 2023, 2.9% of the population had self-reported long COVID according to the Office of National Statistics^5^. Community-based studies report persistent symptoms in approximately 8-17% of cases, with a smaller subset experiencing debilitating impairments^8^. Long COVID symptoms span multiple organ systems and commonly include fatigue, headache, brain fog, dyspnea, anosmia, chest pain, dysautonomia, sleep disorders, with adverse effects including cardiovascular disorders, thrombotic and cerebrovascular disease, type 2 diabetes, and myalgic encephalomyelitis^2,7,9,10^. Overall, more than 200 symptoms have been catalogued^11^.

While some organ damage associated with long COVID may originate during the acute stage of infection, the heterogeneity of long COVID symptoms suggests a complex and incompletely understood pathophysiology^9,12^. Additional proposed contributors to long COVID pathogenesis are viral persistence in tissues, immune dysregulation with or without pathogen reactivation, autoimmunity from molecular mimicry, microvascular and endothelial dysfunction, mitochondrial and metabolic impairments, and autonomic nervous system dysfunction^2,12–14^. Many of these processes overlap with biological pathways implicated in chronic disease development and ageing, including systemic inflammation, metabolic dysregulation, and impaired tissue repair. This overlap raises the possibility that individuals with poorer baseline health or greater physiological burden may be more vulnerable to persistent post-infectious sequelae^8,10^. Shared pathways may include chronic inflammation, endothelial dysfunction, immune dysregulation, and mitochondrial impairment^2,14,15^.

Epidemiological studies consistently show that long COVID risk is associated with older age, one of the strongest predictors^10^, female sex, lower socioeconomic status, obesity, smoking, lack of vaccination, severity of acute COVID-19 infection (requiring hospitalisation or ICU care)^2,16,17^. Poorer pre-pandemic health, with pre-existing comorbidities such as asthma, diabetes and ischemic heart disease, has also been linked to higher risk of developing long COVID, although findings vary across cohorts and diseases^1,2,6–8,10,15,18–22^. Rather than individual conditions acting independently, these associations may reflect the accumulation of health deficits across multiple organ systems. Multimorbidity, the non-random co-occurrence of multiple long-term conditions, captures this cumulative burden. This phenotype already affects over half of adults aged 65 and over in the UK and is projected to impact up to 68% of the UK population by 2035^23^. Importantly, although presentations are heterogeneous, multimorbidity is not randomly distributed: certain diseases consistently cluster together into patterns observed in clinical settings, including neurodegenerative, cardiovascular and metabolic conditions, which may differ in outcomes and healthcare needs^24–28^. These patterns may reflect shared biological mechanisms, environmental exposures, or ageing processes. Emerging evidence suggests that multimorbidity exhibits measurable heritability^29,30^.

Despite growing recognition of long COVID as a multisystem condition, its relationship with pre-existing multimorbidity and potential shared genetic determinants remains poorly understood. Little is known about whether genetic predisposition to multimorbidity influences susceptibility to persistent post-infectious symptoms^10^. Multimorbidity may serve as a meaningful proxy for baseline health status, reflecting cumulative physiological burden across multiple biological systems implicated in post-acute sequelae of viral infection. Preliminary analyses in TwinsUK suggesting an observational association between pre-pandemic multimorbidity clusters and long COVID risk further motivated the present investigation.

Here, we characterise the genetic architecture of multimorbidity patterns in the UK Biobank and investigate whether these patterns share a common genetic basis with long COVID susceptibility. Because both multimorbidity and long COVID are strongly age-associated, older adults represent a particularly relevant population in which to examine shared biological vulnerability^23,27^. Evidence of shared genetic architecture could help identify biological pathways influencing recovery following SARS-CoV-2 infection. Conversely, absence of overlap could suggest that long COVID risk is driven primarily by environmental or post-infectious mechanisms independent of underlying multimorbidity predisposition.

## METHODS

### Study overview

This study followed a multi-step workflow to derive multimorbidity clusters in the UK Biobank, characterise their genetic architecture, and explore their relationship with long COVID. The overall study design, from phenotypic inference and clustering to downstream genetic and causal analyses, is summarised in Figure 1.

**Figure 1:**
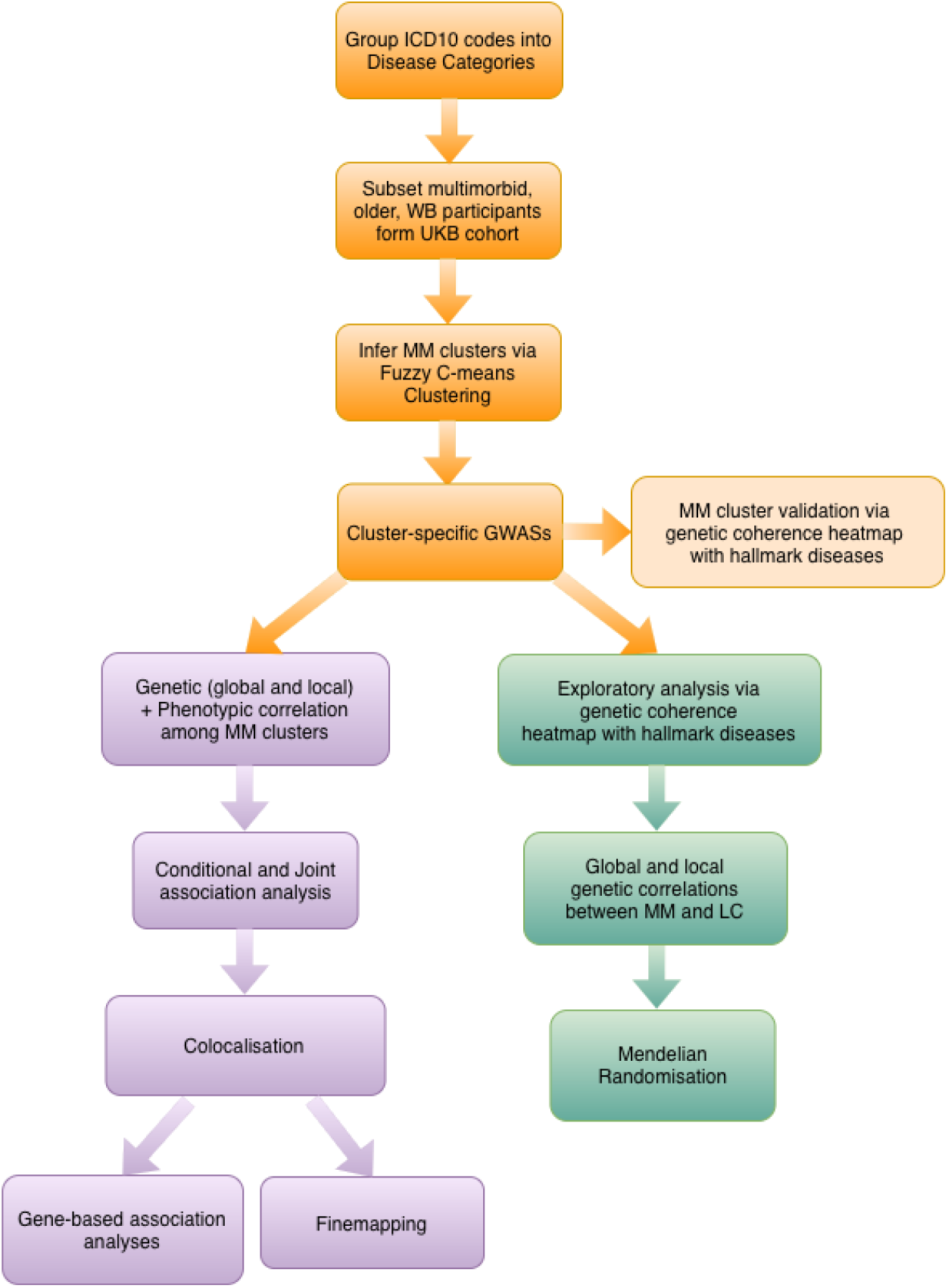
Analytical pipeline Overview of multimorbidity cluster derivation in UK Biobank (UKB), downstream exploration of multimorbidity cluster genetics and investigation of the multimorbidity-long COVID relationship. Clinically-relevant disease categories were based on those published by Calderón-Larrañaga^31^ and derived using ICD10 codes. Multimorbidity participants aged ≥ 65 years and of White British (WB) ancestry were prioritised for analysis. Fuzzy c-means clustering was applied to infer multimorbidity clusters, and cluster membership scores were used as continuous phenotypes in GWAS. A genetic coherence heatmap comparing multimorbidity clusters and hallmark diseases was generated to assess the biological validity of the derived multimorbidity phenotypes. Post-GWAS analyses included cluster-specific SNP-heritability estimation, conditional and joint analysis, inter-cluster genetic and phenotypic correlations, colocalisation, finemapping and gene-based association testing. Long COVID phenotypes were compared to the same hallmark disease set. Shared genetics between long COVID and multimorbidity clusters was explored through global and local genetic correlation. Finally, Mendelian randomisation analyses were performed to investigate the causal effect of multimorbidity on long COVID risk. *Box colours indicate analytical phase: orange, multimorbidity phenotype derivation; purple, genetic characterisation of multimorbidity clusters; green, multimorbidity-long COVID relationship. Blue chevrons indicate key analytical decisions and inputs. GWAS: genome-wide association study; ICD10: International Classification of Diseases, 10th revision; LC: Long Covid; MM: Multimorbiditiy; SNP: single nucleotide polymorphism; UKB: UK Biobank; WB: White British*.

### Phenotype definition and cohort characteristics

To explore the relationship and putative causality between multimorbidity and long COVID, we first inferred groups or clusters of non-randomly co-occurring long-term conditions (Figure 1). We leveraged participant disease data from the UK Biobank (project #90865), a biomedical database containing genetic, demographic, lifestyle and health information from more than 500,000 volunteer participants aged 40 to 69 years, recruited between 2006 and 2010 across the UK^32^. Disease data was extracted from Hospital Episode Statistics, including Hospital inpatient, Death register and Cancer register. Diagnoses were recorded as ICD10 codes from the WHO International Classification of Disease, along with diagnosis date^33^.

To enable interpretable analyses, ICD10 codes designating chronic disease were grouped into clinically relevant disease categories using a previously published classification framework for ageing populations^31^. Of the 60 categories proposed, 59 were retained for our project; with chromosomal abnormalities excluded. Multimorbidity was defined as the presence of at least two disease categories.

Diagnoses were mapped to UK Biobank records. Of 502,371 participants, 394,735 participants had a hospital inpatient record, of whom 323,180 were multimorbid. Participants were then filtered by age at assessment (≥65 years old) and genetic ethnic grouping (White British ancestry), resulting in a final sample of 86,756 participants for clustering and genetic analyses.

### Clustering strategy

Non-random co-occurrence patterns were identified using a clustering framework. We applied fuzzy c-means clustering based on disease categories. Unlike k-means, this soft-clustering strategy allows participants to belong to multiple clusters simultaneously by assigning membership values proportional to the distance between data points and cluster centroids. This relationship is modulated by a fuzzifier variable which controls the degree of overlap between clusters.

Following Vetrano et al., (2020), we applied fuzzy c-means clustering in R using the e1071 package, after dimensionality reduction of the participant-by-disease matrix via multiple correspondence analysis (MCA)^34,35^. Optimal clustering configuration was evaluated across cluster numbers (*K* = 2-12) and fuzzifier values (*m* = 1.1-1.5) using four clustering validity indices: Fukuyama-Sugeno, Calinski-Harabasz, Partition Coefficient, Partition Entropy index. For each fuzzifier value, clustering was repeated 100 times across all tested *K* values to account for randomness in cluster initialisation, and clustering quality indices were averaged to obtain stable performance estimates. Cluster membership values for the selected configuration were used as continuous phenotypes in subsequent genome-wide association studies (GWAS) and are referred to throughout as C1–C6. These phenotypes capture relative similarity of individuals to multimorbidity patterns and may reflect shared biological pathways as well as correlated liability to individual component diseases. The biological validity of the multimorbidity clusters was evaluated using genome-wide genetic correlations with a curated set of “hallmark” diseases in a genetic coherence heatmap, generated using the ComplexHeatmap R package^36^ (Extended Data Figure 3).

### Genetic analysis of multimorbidity clusters

Cluster-specific GWAS of continuous cluster membership scores were performed using the Genome Wide Complex Trait Analysis (GCTA) package, fitting mixed linear models with a sparse genomic relationship matrix to control for sample relatedness and population stratification^37^. Quality control thresholds were set at minor allele frequency >0.01, genotype missingness < 0.02 and imputation INFO score > 0.7^37^. Models were adjusted for age at assessment, sex and the first five genetic principal components. SNP-based heritability of each multimorbidity cluster was estimated using the LD Score Regression package (LDSC)^38,39^. A conditional and joint analysis was then conducted using the GCTA-COJO tool to detect independent genetic association signals characterising the multimorbidity clusters^40^. Prioritised SNPs were functionally annotated.

To explore relationships amongst clusters, pairwise phenotypic correlations were computed and compared with genome-wide genetic correlations estimated with LDSC. Local genetic correlations were estimated using the **L**ocal **A**nalysis of [co]**V**ariant **A**ssociation (LAVA) package^41^ to identify region-specific shared genetic architecture between clusters; summary of results are reported in Supplementary Table 1.

Colocalisation analyses were conducted using the coloc R package to investigate whether regions associated with multiple clusters harboured shared causal signals^42^. Regions surrounding shared independent SNPs were defined as the LD neighbourhood of the lead SNP (r ≥ 0.6), with an additional ±50kb buffer. Evidence for colocalisation was defined as posterior probability of a shared causal variant (PP.H4) > 0.75^42^.

Fine-mapping analyses were performed using FINEMAP (v1.4) to prioritise variants most likely to drive the association within this region^43^. Analyses were conducted within ±1 Mb windows centred on lead variants. Linkage disequilibrium matrices were derived from the UK Biobank reference sample using PLINK. Default FINEMAP settings were used. Evidence for candidate causal variants was assessed using posterior inclusion probabilities (PIPs) and credible sets.

Gene-based association analyses were conducted using MAGMA; detailed results are presented in Supplementary Table 3^44^.

All genetic analyses were performed in the GRCh37 genome build. Analyse were conducted in R version 4.4.1.

### Shared genetics of multimorbidity clusters and long COVID

To investigate shared genetics between multimorbidity and long COVID, GWAS summary statistics for long COVID were obtained from the COVID-19 Host Genetics Initiative (data freeze 4) multi-ancestry discovery meta-analysis, comprising 24 cohorts across 16 countries, totalling 6,450 long COVID cases, 46,208 COVID-19-positive controls and 1,093,955 population controls^45^. Long COVID was defined as “the presence of one or more self-reported COVID-19 symptoms that cannot be explained by an alternative diagnosis, ongoing substantial impact on day-to-day activities, or any diagnosis codes of long COVID”^45^. Four case and control definitions were used in the original study: strict long COVID cases with test-verified SARS-CoV-2, broad long COVID cases including self-reported infection, strict controls with test-confirmed SARS-CoV-2 infection who did not develop long COVID, and broad population controls (ancestry-matched individuals not meeting long COVID criteria, regardless of infection status). For clarity, these phenotypes are referred to in figures and tables as: strictLC_infCtrl, broadLC_infCtrl, strictLC_popCtrl, and broadLC_popCtrl, where *infCtrl* denotes infected controls and *popCtrl* population controls. Although the meta-analysis included multiple ancestries, it was predominantly composed of individuals of European ancestry; therefore, European ancestry LD reference panels were used in downstream analyses. As an exploratory analysis, long COVID phenotypes were compared with the same set of hallmark diseases mentioned previously in pairwise genome-wide genetic correlations (full results in Extended Data Figure 4 and Supplementary Table 4). Analyses were conducted across all four case-control definitions. Comparisons using population controls may conflate long COVID susceptibility with genetic determinants of SARS-CoV-2 infection risk, while infected-only control definitions better isolate long COVID susceptibility but may introduce collider bias. Consistency of findings across definitions therefore provides more robust inference than any single analysis alone.

SNP-based heritability of long COVID phenotypes was obtained from Lammi et al.^45^. Global genetic correlations between long COVID phenotypes and multimorbidity clusters were estimated using LDSC, and local genetic correlations were subsequently estimated with LAVA.

### Causal inference between multimorbidity and long COVID

Potential causal effects of multimorbidity clusters on long COVID risk were examined using Mendelian randomisation (MR), performed with the MRlap R package^46^. MRlap accounts for sample overlap by incorporating cross-trait LDSC intercepts to approximate shared participants between exposure and outcome GWASs, thereby providing unbiased causal effect estimates. This was crucial as the long COVID meta-analysis included UK Biobank participants.

Independent genetic instruments were obtained from multimorbidity cluster GWASs using a genome-wide significance threshold (P < 5 × 10^−8^) and LD-clumping (r^2^ < 0.001, 10 Mb window). Analyses used the HapMap3 SNP set excluding the MHC region. Final MR estimates were derived under the default MRlap parameters, reporting causal effect size and standard error. Clusters with fewer than two independent genome-wide significant instruments were excluded from MR analysis, as reliable bias-corrected causal effect estimation requires a minimum instrument count. An assumption of MR is that multimorbidity-associated variants affect long COVID risk only through their effect on multimorbidity. This assumption may be violated if multimorbidity-associated variants also influence the likelihood of SARS-CoV-2 exposure itself, for example by predicting shielding behaviour, reduced healthcare contact in older multimorbid individuals, or differential survival following acute infection which would introduce bias into causal effect estimates. We note that MRlap does not correct for potential collider bias arising from conditioning on SARS-CoV-2 infection in the long COVID GWAS. MR estimates using population controls mitigate, but do not fully eliminate, the potential collider bias.

## RESULTS

### Clustering of disease domains

The final analytic sample comprised 86,756 multimorbid participants aged ≥65 years (49% female; mean age at assessment 68.1 years), with an average of 6.98 disease categories per individual (Table 1). The prevalence of each disease category in the multimorbid cohort is shown in Extended Data Figure 1.

**Table 1:** Characteristics of UK Biobank participants included in multimorbidity clustering analyses Participants were restricted to individuals of White British ancestry aged ≥65 years with ≥2 disease categories. Healthspan, the period lived free from chronic disease or disability, was approximated here as the age at first recorded diagnosis across disease categories. Values are mean (SD) for continuous variables and n (%) for categorical variables.

| Characteristic | N = 86,756<sup>†</sup> |
| --- | --- |
| Sex |  |
| Female | 42,596 (49%) |
| Male | 44,160 (51%) |
| Mean age at assessment (years) | 68.07 (3.09) |
| Mean healthspan (years) | 64.87 (7.16) |
| Mean number of disease categories | 6.98 (4.40) |
| Mortality | 15,948 (18%) |
| <sup>†</sup> n (%); Mean (SD) |  |

Optimal clustering parameters of the 59 disease categories were selected via visual inspection of four validation indices. The steepest change in clustering quality was observed between *K* = 4 to *K* = 7, indicating a stable range of plausible solutions. Within this range, K = 6 provided a balance between model fit and interpretability while avoiding unnecessary model complexity, yielding clinically meaningful disease groupings consistent with patterns reported in previous multimorbidity studies. We therefore selected K = 6 clusters at m = 1.2 (Extended Data Figure 2).

Disease categories were assigned to clusters based on exclusivity and observed-to-expected prevalence ratios within each cluster. Clusters were named according to their dominant disease pattern: Neurodegenerative (C1), Cardiac (C2), Gastrointestinal (C3), Musculoskeletal (C4), Vascular (C5), and Cancers & Eye disorders (C6) (Figure 2).

**Figure 2:**
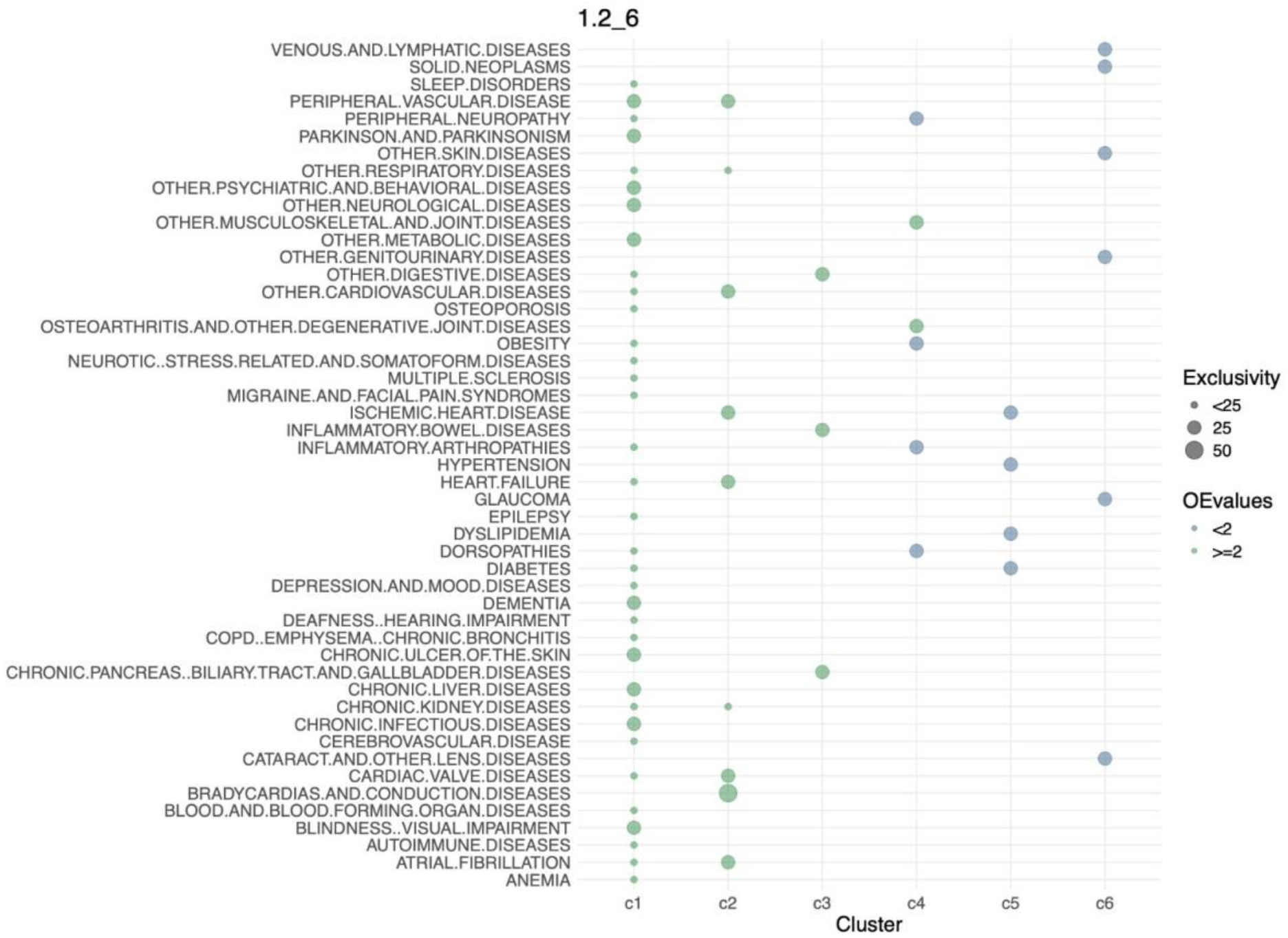
Composition of multimorbidity clusters. Disease categories are shown on the y-axis and multimorbidity clusters on the x-axis. *Exclusivity* indicates how specific a disease is to a given cluster. *OE (Observed-to-Expected)* values quantify the enrichment of each disease in a cluster (compared to its average prevalence across all clusters), adjusted for the size of the cluster and fuzzy membership weights.

Epidemiological characteristics of each multimorbidity cluster are presented in Table 2. Owing to the use of soft clustering, participants exhibit varying degrees of membership across all six clusters.

**Table 2:** Individual multimorbidity cluster characteristics.

| Characteristics | Cluster1 | Cluster2 | Cluster3 | Cluster4 | Cluster5 | Cluster6 |
| --- | --- | --- | --- | --- | --- | --- |
| **NumParticipants<sup>1</sup>** | 7,101 | 8,504 | 11,334 | 14,966 | 16,725 | 28,126 |
| **Female %** | 0.45 | 0.31 | 0.59 | 0.57 | 0.38 | 0.54 |
| **MeanAgeAssessment** | 67.5 | 67.8 | 67.9 | 67.9 | 68.2 | 68.4 |
| **Mortality %** | 0.47 | 0.32 | 0.21 | 0.09 | 0.18 | 0.12 |
| **MeanConditions** | 15.52±4.15 | 11.30±3.55 | 9.72±2.51 | 6.81±2.54 | 5.58±1.93 | 3.35±1.35 |

<sup>1</sup>Number of participants whose highest membership value is in that cluster; this measure is provided for context only and was not used in subsequent analyses.

Clusters differed in disease burden and mortality, with higher proportions of both in neurodegenerative and cardiac profiles and lower mortality rates in lower-burden clusters such as C4 and C6; C5 displayed comparatively elevated mortality despite fewer disease categories, highlighting heterogeneity in risk profiles across clusters.

Clusters demonstrated expected genetic coherence with hallmark diseases (Extended Data Figure 3), supporting biological validity of the inferred phenotypes. We next sought to characterise their genetic architecture through genome-wide association analyses.

### Genome-Wide Association Studies of multimorbidity clusters

Cluster-specific GWAS were conducted in the multimorbid participants of White British ancestry aged 65 years and older (at assessment), treating their cluster membership scores as continuous phenotypes. Manhattan plots summarising associations for SNPs with MAF > 1% across all six phenotypes are shown in Figure 3.

**Figure 3:**
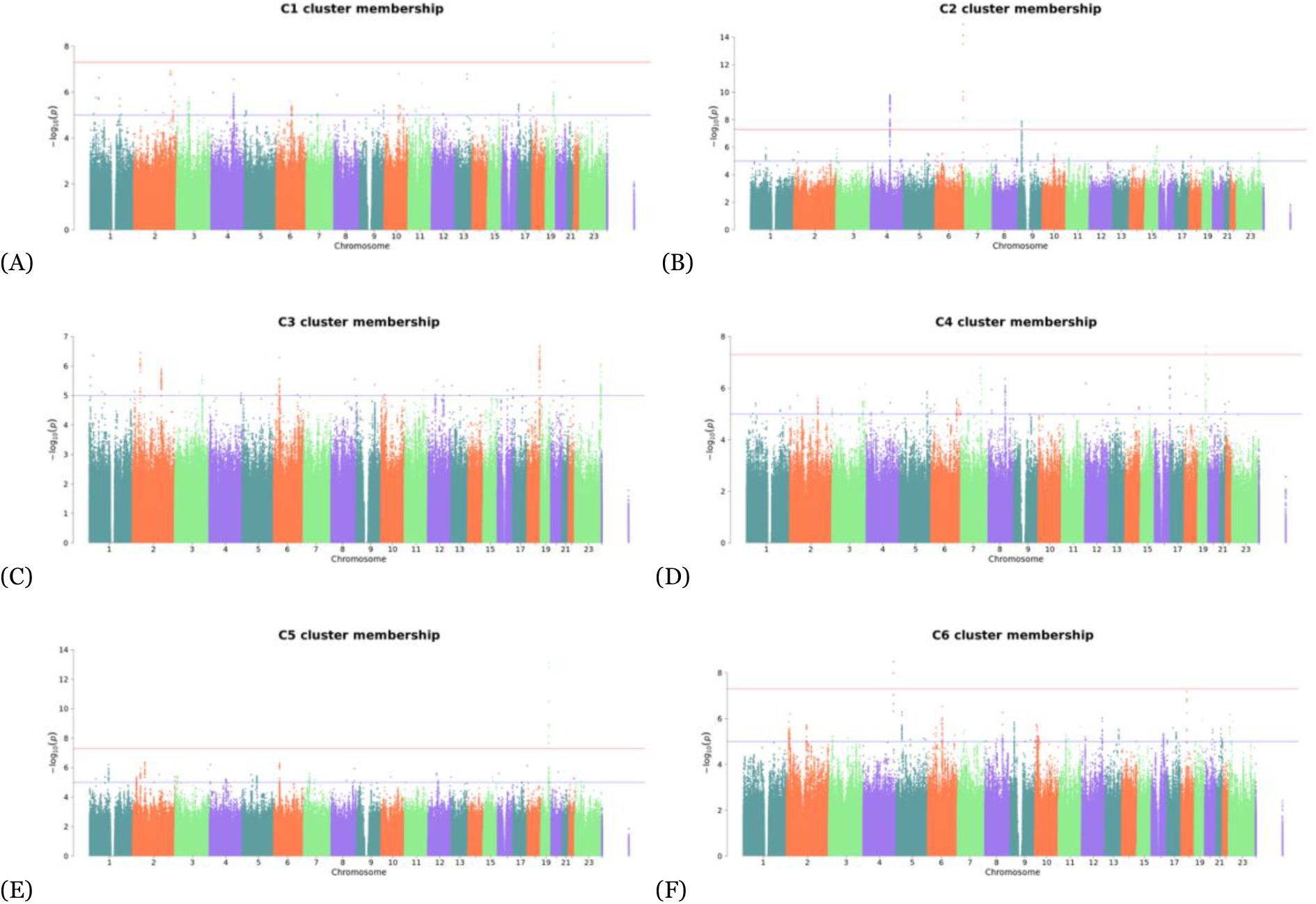
Manhattan plots of genome-wide association results for multimorbidity cluster membership scores. Panels A-F correspond to clusters C1-C6, labelled within each plot. Dots represent individual trait-associated SNPs. The x-axis shows variants’ chromosomal position and the y-axis quantifies the significance of SNP association to the phenotype (displays -log10(p-values)). The red horizontal line represents the genome-wide significant association threshold (P = 5 x 10^−8^) and the blue line the suggestive one (P = 1 x 10^−5^).

Five clusters yielded genome-wide significant associations (*P* < 5 x 10^−8^): 10 loci for C1 (Neurodegenerative), 159 for C2 (Cardiac), 2 for C4 (Musculoskeletal), 11 for C5 (Vascular) and 2 for C6 (Cancers & Eye disorders). No genome-wide significant SNPs were identified for C3 membership (Gastrointestinal), although several suggestive associations were observed (top SNP at *P* = 1.2 x 10^−7^).

SNP-based heritability of cluster membership scores was estimated using LDSC, yielding significant estimates for all six clusters: h^2^_C1_ = 0.039 (SE = 0.006, P = 8.8×10^−12^), h^2^_C2_ = 0.029 (SE = 0.006, P = 2.0×10^−7^), h^2^_C3_ = 0.023 (SE = 0.005, P = 1.7×10^−5^), h^2^_C4_ = 0.029 (SE = 0.006, P = 4.0×10^−7^), h^2^_C5_ = 0.030 (SE = 0.006, P = 2.0×10^−6^), and h^2^_C6_ = 0.052 (SE = 0.006, P = 1.2×10^−16^). GCTA heritability estimates derived from the GWAS mixed model were also obtained; whilst differing in magnitude, cluster ranking was broadly consistent across methods.

To identify the specific independent variants driving these associations, we performed conditional and joint analysis.

### Multimorbidity-associated independent variants

Conditional and joint analysis (COJO) identified six independent genome-wide significant variants associated with multimorbidity cluster membership across four chromosomes (Extended Data Table 1). Negative effect sizes imply alleles associated with weaker cluster membership (i.e., reduced genetic propensity toward that multimorbidity profile).

For C1, a missense variant, **rs429358** (Chr19:45,411,941; T allele; β = −0.0062 ± 0.0010; *P* = 2.6 × 10^−9^), located in *APOE*, where the T allele forms part of the protective ε2 haplotype, was negatively associated with neurodegenerative cluster membership. For C2, three independent variants were identified: **rs72900157** (Chr4:111,710,793; C allele; β = -0.0107 ± 0.0017; *P* = 1.5 × 10^−10^), an intergenic SNP within 4q25 overlapping the lncRNA LINC01438 and near *PITX2*; **rs55730499** (Chr6:161,005,610; C allele; β = -0.0148 ± 0.0018; *P* = 1.2 × 10^−15^), an intronic variant in *LPA*; and **rs10757277** (Chr9:22,124,450; A allele; β = -0.0058 ± 0.0010; *P* = 1.2 × 10^−8^), an intergenic SNP near *CDKN2B-AS1*. Clusters 4 and 5 showed an independent association with a common SNP: **rs1065853** (Chr19:45,413,233; G allele), with opposite effect directions of β = -0.0122 ± 0.0022 (*P* = 2.4 × 10^−8^) for C4 and β = 0.0161 ± 0.0022 (*P* = 7.5 × 10^−14^) for C5, consistent with the negative genome-wide genetic correlation observed between clusters. Rs1065853 lies within the *APOE*-*APOC1* locus. For C6, a single independent signal, **rs71611783** (Chr4:175,315,167; A allele; β = -0.0320 ± 0.0054; *P* = 3.3 × 10^−9^), was identified near *KIAA1712*. No independent signals were detected for C3 (Gastrointestinal disorders).

Beyond individual loci, we next examined the broader genetic relationships amongst clusters.

### Genetic and phenotypic correlation amongst clusters

Genetic and phenotypic correlations amongst clusters were largely consistent, except for the C2-C5 and C1-C4 pairs, which were positively correlated at the genetic level but negatively correlated phenotypically (Figure 4). Overall, nine pairs of clusters demonstrated significant genome-wide genetic correlations, most of which were negative. Strong inverse correlations included: C1 and C6 (*rg* = -0.85; *P* = 1.3 x 10^−40^) and C4 and C5 (*rg* = -0.69; *P* = 6.5 x 10^−9^). Notably, the opposing correlation between C4 and C5 was consistent with the opposing effect directions observed for the shared *APOE– APOC1* locus identified in COJO, supporting a broader inverse genetic relationship between musculoskeletal and vascular multimorbidity profiles. Local genetic correlation analysis using LAVA identified five loci reaching FDR significance across three cluster pairs (C4-C6, C3-C6, and C1-C5), with additional nominally significant loci observed across six further pairs. Effect directions were broadly consistent with genome-wide genetic correlation patterns (Extended Data Table 2 and Supplementary Table 1).

**Figure 4:**
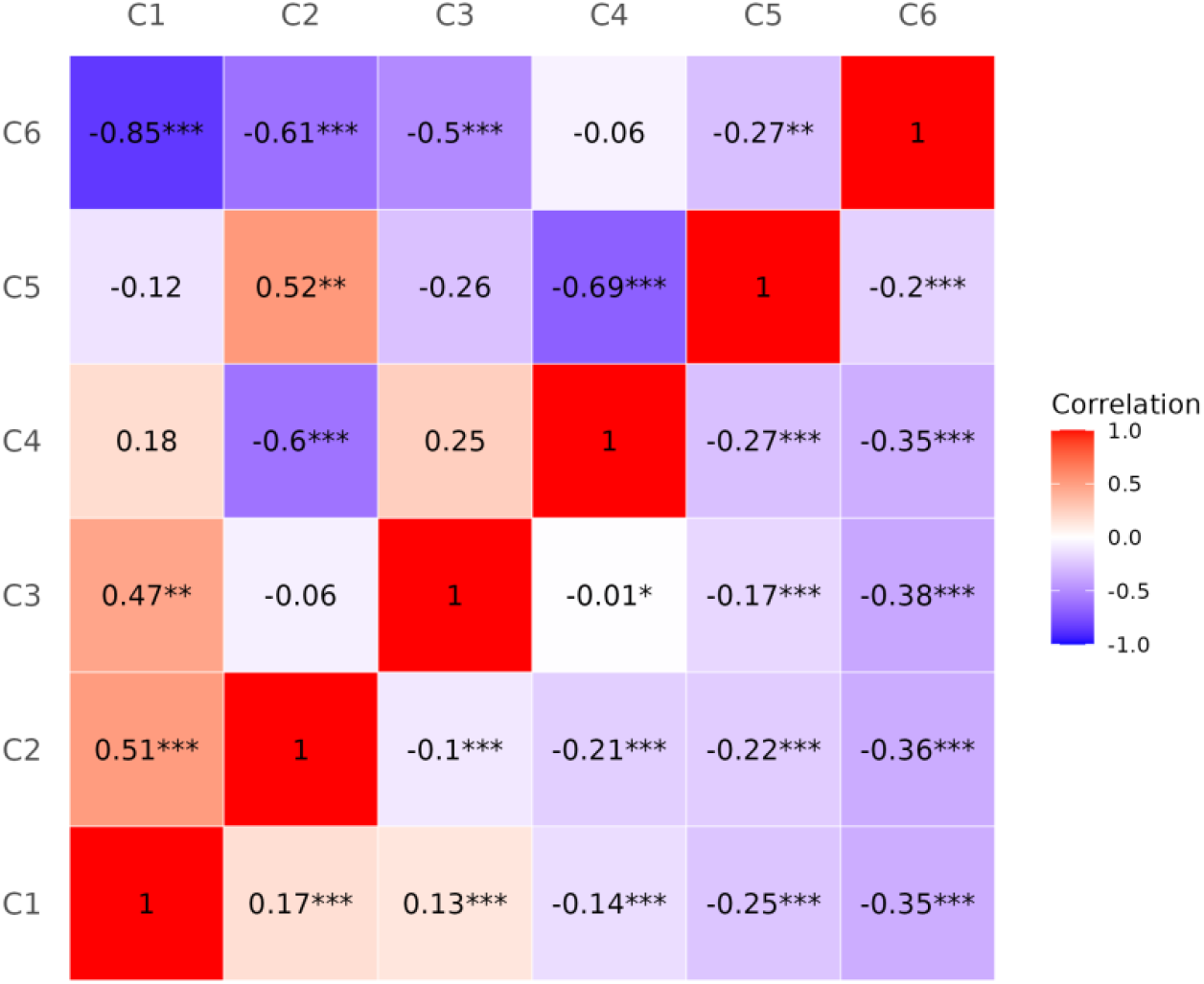
Genetic (upper triangle) and phenotypic (lower triangle) correlations amongst six multimorbidity clusters. Asterisks designate significance level: ***: p < 0.001, **: p < 0.01, *: p < 0.05.

Inter-cluster genetic correlations were consistent with the patterns observed in the cluster-hallmark disease genetic coherence analysis (Figure 4 and Extended Data Figure 3).

To determine whether the shared associations at chromosome 19 reflected a single underlying causal signal, we performed colocalisation analysis.

### Colocalisation

The chromosome 19 region harbouring rs1065853, the common independent SNP for C4 and C5, revealed strong evidence of a shared causal signal (PP.H4=0.999). Variant-level posterior probabilities prioritised rs1065853 (SNP.PP.H4 = 0.79), with residual uncertainty attributable to rs7412 (SNP.PP.H4 = 0.21), a coding variant within the APOE haplotype, which is in strong LD with rs1065853. Both SNPs exhibited consistent opposite effect directions on cluster 4 and 5 memberships (rs1065853: β_C4_= -0.012 and β_C5_ = 0.016; rs7412: β_C4_ = -0.012 and β_C5_ = 0.016); full colocalisation results are provided in Supplementary Table 2.

### Finemapping of the C4/C5 shared locus

Finemapping using FINEMAP of the chromosome 19 locus prioritised a small set of candidate causal variants in strong linkage disequilibrium across both traits, including rs4803743 (PIP = 1.00 in both clusters) and rs1551891 (PIP = 0.76 in cluster 4; PIP = 0.62 in cluster 5). Most shared variants showed opposite directions of effect on cluster membership, consistent with the inverse genetic correlation observed between cluster 4 and 5 memberships. We report the top SNPs of each credible set in Table 3, with posterior inclusion probabilities (PIP) shown for fine-mapped variants shared between clusters C4 and C5; full finemapping results are provided in Extended Data Table 3.

**Table 3:** Finemapped variants shared between C4 and C5 clusters.

| SNP_ID | C4_PIP | C5_PIP | Credible_set(s) | Consequence |
| --- | --- | --- | --- | --- |
| rs4803743 | 1.00 | 1.00 | CS1 | Upstream gene variant of antisense lncRNA (*CTB-171A8.1*, *CEACAM16-AS1*) |
| rs1551891 | 0.76 | 0.62 | CS2 | Upstream gene variant of snoRNA (snoZ6) |
| rs62117161 | 0.24 | 0.08 | CS2 | Upstream gene variant of snoRNA (snoZ6) |
| rs2927487 | 1.00 | 1.00 | CS3 | Downstream gene variant of snoRNA (snoZ6) and upstream gene variant of antisense lncRNA (*CTB-171A8.1*) |
| rs11878852 | 1.00 | 1.00 | CS4 | Intron variant of lncRNA (*CTB-171A8.1*) |

The top credible sets were dominated by non-coding variants mapping to antisense long non-coding RNA and small nucleolar RNA transcripts (*CTB-171A8*.*1, CEACAM16-AS1, snoZ6*) and annotated regulatory elements, suggesting that regulatory mechanisms within the broader *APOE–APOC1* haplotype block mediate this shared causal signal.

Having characterised the genetic architecture of multimorbidity clusters, we next investigated whether these clusters share a common genetic basis with long COVID susceptibility.

### Shared genetics of long COVID and multimorbidity

SNP-heritability of long COVID in the HGI meta-analysis varied markedly across case-control definitions: strict case/infected control: h^2^ = 12.36% (SE = 3.6%), strict case/population control: h^2^ = 1.1% (SE = 1.6%), broad case/infected control: h^2^ = 2.85% (SE = 2.4%), broad case/population control: h^2^ = 0.97% (SE = 0.9%), all on the observed scale ^45^. Only the strict case/infected control definition yielded a significant estimate. The substantially higher heritability of infected-only control definitions compared to population controls is consistent with SARS-CoV-2 infection risk being predominantly environmentally determined; analyses conditioning on confirmed infection therefore better isolate genetic contributions to long COVID susceptibility per se. Differences in age structure between cohorts contributing to multimorbidity and long COVID GWAS may also influence SNP-heritability estimates and reduce power to detect shared genetic architecture, as genetic effects on disease risk can vary across the life course.

Genome-wide genetic correlations between multimorbidity clusters and long COVID phenotypes were not statistically significant (Figure 5; full results in Supplementary Table 5). Directionally, modest positive correlations were observed with the Neurodegenerative (C1) and Musculoskeletal (C4) clusters, whereas inverse correlations were observed with the Vascular (C5) and Cancers & Eye disorders (C6) clusters; however, estimates were imprecise and interpreted cautiously.

**Figure 5:**
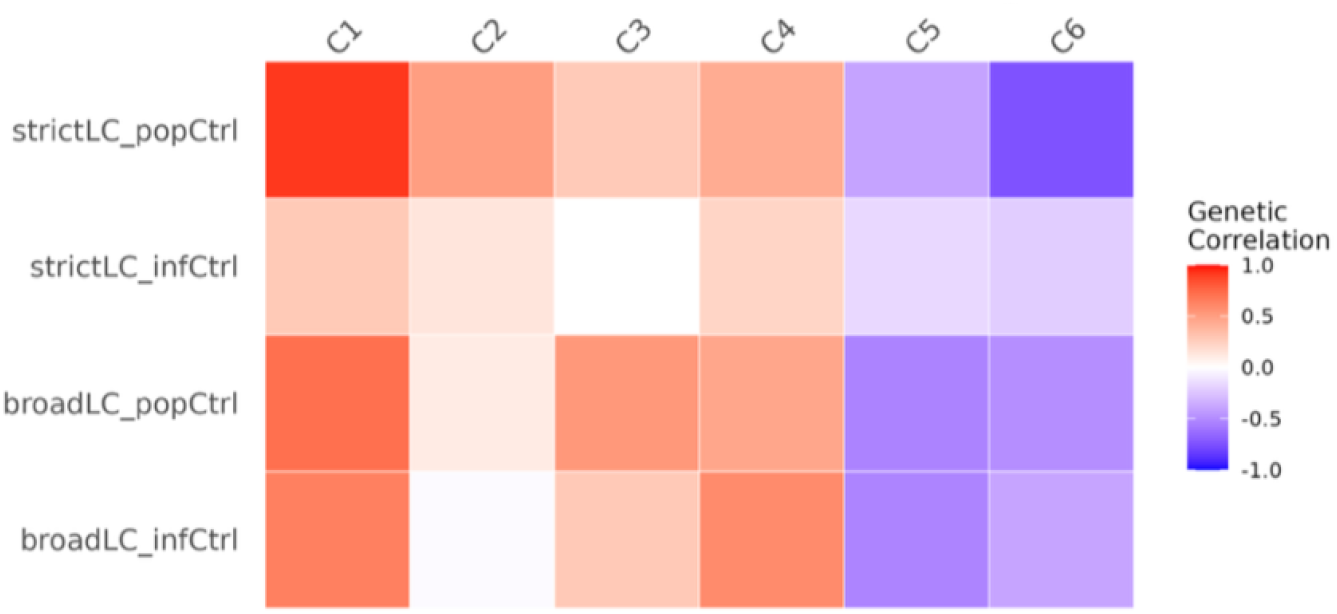
Genome-wide genetic correlations between multimorbidity clusters and long COVID phenotypes

Local genetic correlation analysis with LAVA revealed no loci reaching FDR significance between the long COVID and multimorbidity cluster phenotypes, suggesting that long COVID shares limited detectable common genetic architecture with the multimorbidity clusters at the current sample size (Supplementary Table 6).

These findings are consistent with the absence of significant genome-wide genetic correlations between multimorbidity patterns and long COVID, suggesting that any existent shared architecture is weak or diffuse, rather than driven by discrete loci. To complement these correlation analyses and test for a causal relationship, we performed Mendelian randomisation.

### Mendelian Randomisation

Mendelian randomisation analyses using MRlap, which accounts for sample overlap, weak instrument bias and winner’s curse, was severely limited by instrument availability. Five of six clusters (C1, C3, C4, C5, C6) yielded fewer than two independent genome-wide significant instruments and were therefore excluded from analysis, preventing causal inference for these multimorbidity profiles. Only the cardiac cluster (C2) retained sufficient instruments for MRlap estimation. For C2, no consistent causal effect on long COVID risk was observed: confidence intervals overlapped the null for three of the four long COVID case-control definitions. A nominally positive effect was observed for the broad case-population control long COVID definition (β = 0.0718, SE = 0.0334). However, this was not consistent across alternative definitions and may reflect the broader phenotype structure rather than long COVID susceptibility per se. Given that causal inference was feasible for only one cluster, MR results should be interpreted as hypothesis-generating rather than conclusive.

Extended methods and results are provided in the Supplementary Note.

## DISCUSSION

Multimorbidity is a major and growing public health challenge, particularly in older adults^23^. Here, we investigated its genetic architecture and potential relationship with long COVID. Using fuzzy c-means clustering on 59 clinically derived disease categories, we identified six multimorbidity clusters with distinct clinical and genetic profiles, comprising neurodegenerative, cardiac, gastrointestinal, musculoskeletal, vascular, and cancers and eye disorders domains. Genome-wide and locus-specific analyses, including comparing clusters to a set of hallmark diseases, supported the biological relevance of these phenotypes and highlighted both shared and antagonistic genetic architecture across multimorbidity patterns.

All six multimorbidity clusters showed significant SNP-heritability and five yielded genome-wide significant loci, demonstrating that cluster membership scores capture genuinely heritable phenotypes with genetic architecture extending beyond single disease effects. Because these scores reflect patterns of disease co-occurrence, associated loci likely implicate both shared biological mechanisms within clusters and genetic liability to strongly contributing component diseases. Effect sizes were modest, consistent with the highly polygenic architecture expected for composite multimorbidity phenotypes. The largest signal count was observed for the Cardiac cluster (C2), with loci at *LPA, PITX2* and *CDKN2B-AS1* replicating known cardiometabolic associations. *APOE*-linked signals were associated with the Neurodegenerative (C1), Musculoskeletal (C4), and Vascular disorders (C5) clusters, with opposite directions of effect consistent with pleiotropic influences of *APOE* on lipid metabolism, neuroinflammation, and metabolic regulation. As expected for polygenic traits, effect sizes were small, reflecting the modest contribution of individual common variants to complex multimorbidity phenotypes.

Genome-wide genetic correlation analyses further supported opposing relationships across clusters, particularly between Neurodegenerative (C1) or Cardiac (C2) clusters and the Cancer & Eye disorders cluster (C6), as well as between the Musculoskeletal (C4) and Vascular disorders (C5) clusters. While negative genetic correlations do not necessarily imply direct biological antagonism, but may reflect distinct genetic architectures influencing liability to different disease patterns. Furthermore, such inverse relationships are consistent with epidemiological observations, including the well-studied inverse comorbidity between neurodegenerative diseases and risk for several cancers. Alzheimer’s and Parkinson’s diseases, for example, are associated with a lower risk of many malignancies such as lung, colorectal, and prostate cancers. Conversely, higher cancer susceptibility has been linked to reduced risk of neurodegeneration^47–49.^ These patterns are thought to reflect differences in regulation of cell proliferation and apoptosis, oxidative stress responses and pathways such as *p53* and *PIN1* that promote either cell survival or programmed death^47–49^.

At the locus-level, most inverse relationships between clusters appeared to be mediated by distinct genetic effects acting within shared regions or biological systems rather than single variants acting in opposite directions. A notable exception was identified at the chromosome 19 *APOE* locus, where conditional and joint analysis and colocalisation supported a shared signal between the Musculoskeletal (C4) and Vascular disorders (C5) clusters. Effect estimates of the lead variant rs1065853 showed opposing directions of association between clusters (negative for C4 and positive for C5), consistent with their negative global genetic correlation. Remaining posterior probability from colocalisation was assigned to rs7412, a well characterised coding variant within the APOE haplotype associated with Alzheimer’s disease risk, which also demonstrated consistent effect directions across these clusters. While rs1065853 was intergenic, both variants map to the *APOE-APOC1* region, a locus with established roles in lipid metabolism, immune function, and systemic inflammation. Finemapping independently prioritised rs4803743 as the most probable shared causal variant for both clusters, which also demonstrated opposite directions of effect between C4 and C5, although with the reverse orientation relative to rs1065853 and rs7412. Notably, rs4803743 was not in strong linkage disequilibrium with rs1065853, suggesting that multiple variants within the *APOE–APOC1* region may contribute to antagonistic multimorbidity profiles. Together, these findings support a complex locus architecture underlying the inverse relationship between musculoskeletal and vascular disease clusters.

These findings highlight the complexity of the genetic architecture underlying inverse relationships between multimorbidity clusters (inverse comorbidity). While most antagonistic relationships between multimorbidity clusters appear to be mediated by distinct variants or mechanisms within shared biological pathways, specific loci such as *APOE* may contain multiple variants with distinct effects that contribute to opposing disease profiles across clusters.

Despite robust genetic signal within multimorbidity clusters, we observed limited evidence of shared genetic architecture with long COVID. Genome-wide genetic correlations between long COVID and all clusters were uniformly non-significant, with imprecise directional trends only; power may have been further reduced by the use of multi-ancestry summary statistics. Similarly, correlations between long COVID and hallmark diseases were not significant and varied across case definitions, reflecting definitional heterogeneity and limited statistical power^8^ (Supplementary Table 4; Extended Data Figure 4). Together with the variable and largely modest SNP-heritability of long COVID phenotypes-significant only under the strictest case-control definition-these results suggest that susceptibility to long COVID is unlikely to be strongly shaped by the inherited genetic architecture underlying multimorbidity patterns in this sample. This is consistent with previous studies suggesting that long COVID is influenced by heterogeneous environmental, immunological, and clinical factors, which may dilute detectable common variant heritability^45^. Genetic studies of long COVID remain limited and have yielded modest and phenotype-dependent evidence for common variant contribution. Recent work has also highlighted challenges in replicating long COVID genetic associations across cohorts, with evidence suggesting a broader but heterogeneous genetic signal that may not be well captured by conventional GWAS approaches^50^. Mendelian randomisation analyses offered only limited additional insight. Instrument scarcity restricted causal inference to C2, and no consistent causal effect of cardiac multimorbidity on long COVID risk was identified across long COVID definitions.

Taken together, the convergent null findings from genetic correlation and MR corroborate epidemiological studies reporting inconsistent associations between cardiometabolic conditions and long COVID risk^8^ and suggest that long COVID risk may be shaped by other factors rather than by polygenic burden underlying multimorbidity. While larger cohorts may refine these relationships, our results indicate that genetic predisposition to multimorbidity does not strongly influence long COVID susceptibility at the population level.

Several limitations should be considered. First, the UK Biobank cohort is not fully representative of the UK population: participants are generally healthier, wealthier, and less ethnically diverse, which may limit generalisability^51^. Second, analyses were restricted to White British participants to minimise population stratification, hindering applicability to other ancestries. Third, the multimorbidity classification was developed for older populations, and conditions such as autism and ADHD, which are not age-related in onset, may be under-ascertained in older cohorts and were therefore excluded. Findings may not extend to younger individuals. Fourth, cluster structure and annotation were partly determined by parameter selection and visual interpretation; alternative specifications could yield different profiles. Fifth, heterogeneity in long COVID definitions and low statistical power complicate interpretation of results, as different case-control specifications capture distinct aspects of susceptibility. Sixth, long COVID GWAS analyses based on infected-only controls condition on SARS-CoV-2 infection, which may introduce collider bias if genetic factors associated with multimorbidity also influence infection risk or exposure probability^52^. Analyses using population controls partially mitigate this limitation. Finally, multimorbidity cluster GWASs were restricted to UK Biobank participants aged ≥65 years and of White British ancestry, whereas the HGI long COVID meta-analysis included multiple ancestries, with mean case ages ranging from 29 to 64 years, with most cohorts in the 48–64 year range. SNP-heritability estimates and genetic correlations may vary across the life course. Genetic instruments derived in older multimorbid adults may therefore capture biological processes less strongly represented in the younger long COVID GWAS population, potentially attenuating our ability to detect shared genetic architecture between the two conditions.

## CONCLUSIONS

This work characterised the genetic architecture of multimorbidity patterns in older adults and their relationship to long COVID. The identified clusters show strong internal coherence and biologically plausible genetic signatures, reinforcing that these phenotypes reflect structured, partly heritable patterns. The absence of clear shared genetic architecture with long COVID suggests that predisposition to long COVID may be shaped more by acute infection responses, immune and viral persistence mechanisms, or non-genetic factors than by genetic predisposition to multimorbidity. As better powered and more precisely defined long COVID GWAS become available, integrating these with multimorbidity architectures across ancestries and age groups may clarify whether certain multimorbidity trajectories contribute to long COVID susceptibility or recovery outcomes.

## Data Availability

All data produced in the present study are contained in the manuscript, extended data, and supplementary files, or are available upon reasonable request to the authors.

## ACKNOWLEDGEMENTS

This research was conducted using the UK Biobank Resource under Application Number 90865. The authors thank the Department of Twin Research, Ageing and Genetic Epidemiology at St Thomas’ Hospital, London.

## FUNDING

This work was supported by the CONVALESCENCE study (NIHR #COV-LT-0009) and by the Chronic Disease Research Foundation.

NO COMPETING INTERESTS

EXTENDED AND SUPPLEMENTAL DATA available

