## Supplemental Data for "Exploring the genetic architecture of multimorbidity and its impact on long COVID risk"

### Supplementary Table 1. Bivariate local genetic correlation summary amongst multimorbidity clusters

| **pair** | **n_bivar** | **n_nominal_0.05** | **n_FDR_pair_0.05** | **min_p** | **top_chr** | **top_start** | **top_stop** | **top_rho_ci** |
| --- | --- | --- | --- | --- | --- | --- | --- | --- |
| **C4 ~ C6** | 4 | 1 | 1 | 0.003 | 3 | 68319771 | 69784969 | -0.954 (-1.000, -0.495) |
| **C3 ~ C6** | 4 | 3 | 3 | 0.01 | 2 | 65938003 | 67224027 | -1.000 (-1.000, -0.390) |
| **C2 ~ C6** | 5 | 3 | 0 | 0.01 | 12 | 82576130 | 83551125 | -0.984 (-1.000, -0.414) |
| **C3 ~ C5** | 5 | 3 | 0 | 0.02 | 6 | 32629240 | 32682213 | -1.000 (-1.000, -0.342) |
| **C1 ~ C6** | 6 | 1 | 0 | 0.02 | 7 | 105312564 | 106399739 | -0.857 (-1.000, -0.309) |
| **C2 ~ C5** | 3 | 1 | 0 | 0.02 | 14 | 95946793 | 97174314 | -0.847 (-1.000, -0.225) |
| **C1 ~ C5** | 1 | 1 | 1 | 0.03 | 6 | 91097025 | 92015279 | -0.855 (-1.000, -0.160) |
| **C4 ~ C5** | 2 | 0 | 0 | 0.06 | 5 | 154599751 | 155459556 | -0.682 (-1.000, 0.005) |
| **C2 ~ C4** | 2 | 0 | 0 | 0.15 | 3 | 2140012 | 3128152 | -0.632 (-1.000, 0.286) |
| **C1 ~ C2** | 4 | 0 | 0 | 0.26 | 5 | 97316602 | 98567769 | 0.437 (-0.447, 1.000) |
| **C3 ~ C4** | 3 | 0 | 0 | 0.26 | 2 | 135160198 | 137061003 | -0.440 (-1.000, 0.429) |
| **C1 ~ C3** | 2 | 0 | 0 | 0.55 | 6 | 66445496 | 67036787 | 0.235 (-0.832, 1.000) |
| **C2 ~ C3** | 2 | 0 | 0 | 0.72 | 6 | 143083348 | 144791455 | -0.144 (-1.000, 1.000) |

Supplementary Table 2. Colocalisation results for LAVA-significant multimorbidity cluster membership loci. Table headers are: traits: MM cluster phenotypes being compared; region: genomic location test; nsnps: number of SNPs included in test, H0 to H4: posterior probabilities for each of the five coloc hypotheses, where H0: no association between compared traits, H1: association to trait 1 only, H2: association to trait 2 only, H3: association to both traits, distinct causal variants, H4: association to both traits, shared causal variants.

| **traits** | **region** | **nsnps** | **PP.H0.abf** | **PP.H1.abf** | **PP.H2.abf** | **PP.H3.abf** | **PP.H4.abf** |
| --- | --- | --- | --- | --- | --- | --- | --- |
| **C1 ~ C5** | chr6_91097025_92015279 | 4680 | 0.69 | 0.12 | 0.15 | 0.03 | 0.01 |
| **C1 ~ C6** | chr7_105312564_106399739 | 6195 | 0.09 | 0.03 | 0.50 | 0.15 | 0.23 |
| **C2 ~ C5** | chr14_95946793_97174314 | 7486 | 0.59 | 0.18 | 0.17 | 0.05 | 0.01 |
| **C2 ~ C6** | chr12_82576130_83551125 | 4573 | 0.73 | 0.12 | 0.12 | 0.02 | 0.01 |
| **C2 ~ C6** | chr7_19131027_20122179 | 7435 | 0.50 | 0.22 | 0.16 | 0.07 | 0.05 |
| **C2 ~ C6** | chr9_36749274_37687112 | 4940 | 0.55 | 0.09 | 0.28 | 0.05 | 0.02 |
| **C3 ~ C5** | chr6_32539568_32586784 | 2770 | 0.65 | 0.04 | 0.27 | 0.02 | 0.02 |
| **C3 ~ C5** | chr6_32586785_32629239 | 3350 | 0.19 | 0.41 | 0.08 | 0.16 | 0.16 |
| **C3 ~ C5** | chr6_32629240_32682213 | 3222 | 0.38 | 0.11 | 0.32 | 0.09 | 0.11 |
| **C3 ~ C6** | chr2_65938003_67224027 | 6812 | 0.40 | 0.23 | 0.12 | 0.07 | 0.18 |
| **C3 ~ C6** | chr6_32897999_33194975 | 4036 | 0.19 | 0.61 | 0.03 | 0.10 | 0.08 |
| **C3 ~ C6** | chr6_66445496_67036787 | 4111 | 0.74 | 0.11 | 0.11 | 0.02 | 0.02 |
| **C4 ~ C6** | chr3_68319771_69784969 | 8518 | 0.56 | 0.18 | 0.18 | 0.06 | 0.01 |

Note: In LAVA-significant regions, colocalisation analyses demonstrated weak support for shared causal variants, with most pairs tested showing PP.H4 ≤ 0.05.

Supplementary Table 3: MAGMA gene-based summary results table. Only FDR-significant genes are featured.

| **SYMBOL** | **NSNPS** | **N** | **ZSTAT** | **P** | **P_bonf** | **P_fdr** | **CHR** | **START** | **STOP** | **STRAND** |
| --- | --- | --- | --- | --- | --- | --- | --- | --- | --- | --- |
| **C1** | | | | | | | | | | |
| **ECEL1** | 51 | 86000 | 4.5202 | 3.1E+06 | 0.06 | 0.019 | 2 | 233344537 | 233352532 | - |
| **APOE** | 12 | 86000 | 5.5474 | 1.5E-08 | 0.0003 | 0.0003 | 19 | 45409039 | 45412650 | + |
| **APOC1** | 19 | 86000 | 5.0191 | 2.6E-07 | 0.005 | 0.002 | 19 | 45417577 | 45422606 | + |
| **C4** | | | | | | | | | | |
| **CCDC33** | 516 | 86000 | 4.5714 | 2.4E-06 | 0.04 | 0.02 | 15 | 74528630 | 74628813 | + |
| **APOE** | 12 | 86000 | 4.5793 | 2.3E-06 | 0.04 | 0.02 | 19 | 45409039 | 45412650 | + |
| **C5** | | | | | | | | | | |
| **APOE** | 12 | 86000 | 5.2341 | 8.3E-08 | 0.002 | 0.001521 | 19 | 45409039 | 45412650 | + |

Note: MAGMA identified FDR-significant genes for three clusters, with APOE significant across multiple clusters. Despite limited evidence for shared causal variants at the SNP level across most regions, the cross-cluster association of APOE suggests convergence of genetic effects at the gene level, potentially mediated by distinct variants within the APOE locus. Specifically, three genes were identified for Cluster 1 membership including APOE and APOC1 on chromosome 19 and ECEL1 on chromosome 2. For Cluster 4 membership, CCDC33 on chromosome 15 and APOE remained significant after FDR correction. For Cluster 5 membership, APOE was the only gene reported as FDR-significant.

Supplementary Table 4. Genome-wide genetic correlations between long COVID and multimorbidity clusters VS hallmark diseases. Abbreviations are IHD: Ischemic Heart Disease, Systolic HF: Systolic heart failure, IBD: Inflammatory Bowel Disease, Prostate carci: prostate carcinoma, Periph neuro: peripheral neuropathy.

| **Trait1** | **Trait2** | **rg** | **se** | **z** | **p** |
| --- | --- | --- | --- | --- | --- |
| C1 | Aortic_stenosis | **0.26** | 0.07 | 3.61 | 0 |
| C2 | Aortic_stenosis | **0.45** | 0.07 | 6.01 | 0 |
| C3 | Aortic_stenosis | **0.05** | 0.08 | 0.56 | 0.57 |
| C4 | Aortic_stenosis | **-0.13** | 0.08 | -1.69 | 0.09 |
| C5 | Aortic_stenosis | **0.20** | 0.08 | 2.41 | 0.02 |
| C6 | Aortic_stenosis | **-0.38** | 0.06 | -6.41 | 0 |
| broadLC_popCtrl | Aortic_stenosis | **0.04** | 0.20 | 0.21 | 0.83 |
| broadLC_infCtrl | Aortic_stenosis | **0.09** | 0.19 | 0.46 | 0.65 |
| strictLC_popCtrl | Aortic_stenosis | **0.25** | 0.33 | 0.77 | 0.44 |
| strictLC_infCtrl | Aortic_stenosis | **0.12** | 0.11 | 1.11 | 0.27 |
| C1 | Bradycardia | **0.29** | 0.38 | 0.78 | 0.43 |
| C2 | Bradycardia | **0.37** | 0.39 | 0.97 | 0.33 |
| C3 | Bradycardia | **0.13** | 0.35 | 0.36 | 0.72 |
| C4 | Bradycardia | **-0.28** | 0.36 | -0.77 | 0.44 |
| C5 | Bradycardia | **-0.06** | 0.33 | -0.19 | 0.85 |
| C6 | Bradycardia | **-0.13** | 0.25 | -0.55 | 0.58 |
| broadLC_popCtrl | Bradycardia | **0.32** | 0.63 | 0.50 | 0.62 |
| broadLC_infCtrl | Bradycardia | **0.11** | 0.56 | 0.20 | 0.85 |
| strictLC_popCtrl | Bradycardia | **0.05** | 0.69 | 0.08 | 0.94 |
| strictLC_infCtrl | Bradycardia | **-0.29** | 0.39 | -0.74 | 0.46 |
| C1 | Chronic_pancreatitis | **0.27** | 0.15 | 1.84 | 0.07 |
| C2 | Chronic_pancreatitis | **-0.03** | 0.17 | -0.18 | 0.85 |
| C3 | Chronic_pancreatitis | **0.49** | 0.21 | 2.37 | 0.02 |
| C4 | Chronic_pancreatitis | **0.21** | 0.15 | 1.37 | 0.17 |
| C5 | Chronic_pancreatitis | **-0.10** | 0.17 | -0.59 | 0.55 |
| C6 | Chronic_pancreatitis | **-0.33** | 0.13 | -2.44 | 0.01 |
| broadLC_popCtrl | Chronic_pancreatitis | **0.55** | 0.47 | 1.17 | 0.24 |
| broadLC_infCtrl | Chronic_pancreatitis | **0.68** | 0.47 | 1.43 | 0.15 |
| strictLC_popCtrl | Chronic_pancreatitis | **0.40** | 0.49 | 0.82 | 0.42 |
| strictLC_infCtrl | Chronic_pancreatitis | **0.30** | 0.21 | 1.43 | 0.15 |
| C1 | Coeliac | **0.25** | 0.11 | 2.21 | 0.03 |
| C2 | Coeliac | **0.15** | 0.15 | 0.96 | 0.34 |
| C3 | Coeliac | **0.20** | 0.16 | 1.30 | 0.19 |
| C4 | Coeliac | **-0.02** | 0.15 | -0.15 | 0.88 |
| C5 | Coeliac | **-0.30** | 0.14 | -2.19 | 0.03 |
| C6 | Coeliac | **-0.07** | 0.11 | -0.58 | 0.56 |
| broadLC_popCtrl | Coeliac | **-0.01** | 0.30 | -0.03 | 0.97 |
| broadLC_infCtrl | Coeliac | **-0.09** | 0.29 | -0.31 | 0.75 |
| strictLC_popCtrl | Coeliac | **-0.01** | 0.38 | -0.04 | 0.97 |
| strictLC_infCtrl | Coeliac | **-0.09** | 0.18 | -0.50 | 0.62 |
| C1 | Dementia | **0.45** | 0.18 | 2.55 | 0.01 |
| C2 | Dementia | **0.45** | 0.18 | 2.45 | 0.01 |
| C3 | Dementia | **0.06** | 0.21 | 0.31 | 0.76 |
| C4 | Dementia | **0.27** | 0.20 | 1.33 | 0.18 |
| C5 | Dementia | **-0.15** | 0.19 | -0.80 | 0.42 |
| C6 | Dementia | **-0.47** | 0.16 | -2.91 | 0 |
| broadLC_popCtrl | Dementia | **-0.12** | 0.33 | -0.36 | 0.72 |
| broadLC_infCtrl | Dementia | **-0.25** | 0.37 | -0.67 | 0.50 |
| strictLC_popCtrl | Dementia | **0.08** | 0.49 | 0.17 | 0.87 |
| strictLC_infCtrl | Dementia | **-0.01** | 0.22 | -0.04 | 0.96 |
| C1 | Diabetes | **0.65** | 0.06 | 10.22 | 0 |
| C2 | Diabetes | **0.41** | 0.08 | 5.25 | 0 |
| C3 | Diabetes | **0.42** | 0.08 | 5.56 | 0 |
| C4 | Diabetes | **0** | 0.07 | 0.01 | 1 |
| C5 | Diabetes | **0.15** | 0.07 | 2.24 | 0.03 |
| C6 | Diabetes | **-0.69** | 0.05 | -13.15 | 0 |
| broadLC_popCtrl | Diabetes | **0.23** | 0.19 | 1.18 | 0.24 |
| broadLC_infCtrl | Diabetes | **0.27** | 0.19 | 1.43 | 0.15 |
| strictLC_popCtrl | Diabetes | **0.49** | 0.39 | 1.25 | 0.21 |
| strictLC_infCtrl | Diabetes | **0.20** | 0.08 | 2.42 | 0.02 |
| C1 | Glaucoma | **0.11** | 0.07 | 1.70 | 0.09 |
| C2 | Glaucoma | **0.04** | 0.07 | 0.54 | 0.59 |
| C3 | Glaucoma | **-0.02** | 0.08 | -0.20 | 0.84 |
| C4 | Glaucoma | **0.08** | 0.07 | 1.05 | 0.29 |
| C5 | Glaucoma | **0.03** | 0.08 | 0.34 | 0.73 |
| C6 | Glaucoma | **-0.11** | 0.05 | -2.12 | 0.03 |
| broadLC_popCtrl | Glaucoma | **0.03** | 0.13 | 0.21 | 0.83 |
| broadLC_infCtrl | Glaucoma | **0.04** | 0.13 | 0.29 | 0.77 |
| strictLC_popCtrl | Glaucoma | **0.26** | 0.27 | 0.99 | 0.32 |
| strictLC_infCtrl | Glaucoma | **0.11** | 0.09 | 1.30 | 0.19 |
| C1 | Hyperlipidemia | **0.43** | 0.07 | 6.10 | 0 |
| C2 | Hyperlipidemia | **0.27** | 0.07 | 3.73 | 0 |
| C3 | Hyperlipidemia | **0.27** | 0.09 | 2.93 | 0 |
| C4 | Hyperlipidemia | **0.03** | 0.09 | 0.38 | 0.71 |
| C5 | Hyperlipidemia | **0.39** | 0.08 | 4.62 | 0 |
| C6 | Hyperlipidemia | **-0.63** | 0.06 | -9.77 | 0 |
| broadLC_popCtrl | Hyperlipidemia | **0.13** | 0.19 | 0.70 | 0.48 |
| broadLC_infCtrl | Hyperlipidemia | **0.17** | 0.19 | 0.93 | 0.35 |
| strictLC_popCtrl | Hyperlipidemia | **0.39** | 0.33 | 1.17 | 0.24 |
| strictLC_infCtrl | Hyperlipidemia | **0.15** | 0.10 | 1.56 | 0.12 |
| C1 | Hypertension | **0.36** | 0.05 | 7.85 | 0 |
| C2 | Hypertension | **0.52** | 0.07 | 7.94 | 0 |
| C3 | Hypertension | **0.10** | 0.06 | 1.50 | 0.13 |
| C4 | Hypertension | **-0.06** | 0.05 | -1.31 | 0.19 |
| C5 | Hypertension | **0.52** | 0.06 | 8.75 | 0 |
| C6 | Hypertension | **-0.68** | 0.05 | -14.58 | 0 |
| broadLC_popCtrl | Hypertension | **-0.04** | 0.12 | -0.34 | 0.74 |
| broadLC_infCtrl | Hypertension | **-0.05** | 0.11 | -0.47 | 0.64 |
| strictLC_popCtrl | Hypertension | **0.05** | 0.13 | 0.40 | 0.69 |
| strictLC_infCtrl | Hypertension | **0.03** | 0.06 | 0.49 | 0.63 |
| C1 | IBD | **0.05** | 0.08 | 0.57 | 0.57 |
| C2 | IBD | **0.12** | 0.10 | 1.11 | 0.27 |
| C3 | IBD | **0.19** | 0.12 | 1.56 | 0.12 |
| C4 | IBD | **-0.14** | 0.11 | -1.28 | 0.20 |
| C5 | IBD | **-0.04** | 0.10 | -0.37 | 0.71 |
| C6 | IBD | **-0.05** | 0.08 | -0.58 | 0.56 |
| broadLC_popCtrl | IBD | **0.30** | 0.27 | 1.10 | 0.27 |
| broadLC_infCtrl | IBD | **0.41** | 0.28 | 1.45 | 0.15 |
| strictLC_popCtrl | IBD | **0.30** | 0.34 | 0.87 | 0.39 |
| strictLC_infCtrl | IBD | **0.12** | 0.12 | 1.01 | 0.31 |
| C1 | IHD | **0.70** | 0.06 | 10.86 | 0 |
| C2 | IHD | **0.83** | 0.08 | 10.53 | 0 |
| C3 | IHD | **0.30** | 0.08 | 3.96 | 0 |
| C4 | IHD | **-0.22** | 0.07 | -3.31 | 0 |
| C5 | IHD | **0.37** | 0.07 | 5.53 | 0 |
| C6 | IHD | **-0.86** | 0.05 | -17.37 | 0 |
| broadLC_popCtrl | IHD | **0.14** | 0.15 | 0.95 | 0.34 |
| broadLC_infCtrl | IHD | **0.11** | 0.15 | 0.74 | 0.46 |
| strictLC_popCtrl | IHD | **0.47** | 0.41 | 1.16 | 0.25 |
| strictLC_infCtrl | IHD | **0.14** | 0.08 | 1.80 | 0.07 |
| C1 | Osteoarthritis | **0.28** | 0.36 | 0.78 | 0.43 |
| C2 | Osteoarthritis | **-0.01** | 0.35 | -0.02 | 0.98 |
| C3 | Osteoarthritis | **0.42** | 0.46 | 0.92 | 0.36 |
| C4 | Osteoarthritis | **0.24** | 0.30 | 0.80 | 0.42 |
| C5 | Osteoarthritis | **-0.28** | 0.35 | -0.79 | 0.43 |
| C6 | Osteoarthritis | **-0.24** | 0.30 | -0.80 | 0.42 |
| broadLC_popCtrl | Osteoarthritis | **1.31** | 1.23 | 1.06 | 0.29 |
| broadLC_infCtrl | Osteoarthritis | **1.23** | 1.12 | 1.10 | 0.27 |
| strictLC_popCtrl | Osteoarthritis | **-0.45** | 1.19 | -0.37 | 0.71 |
| strictLC_infCtrl | Osteoarthritis | **-0.36** | 0.56 | -0.65 | 0.51 |
| C1 | Parkinsons | **0** | 0.07 | 0.05 | 0.96 |
| C2 | Parkinsons | **-0.15** | 0.08 | -1.84 | 0.07 |
| C3 | Parkinsons | **0.06** | 0.09 | 0.68 | 0.50 |
| C4 | Parkinsons | **0.04** | 0.08 | 0.43 | 0.67 |
| C5 | Parkinsons | **0.02** | 0.08 | 0.25 | 0.80 |
| C6 | Parkinsons | **0.02** | 0.06 | 0.37 | 0.71 |
| broadLC_popCtrl | Parkinsons | **0.08** | 0.18 | 0.46 | 0.65 |
| broadLC_infCtrl | Parkinsons | **0.07** | 0.18 | 0.38 | 0.70 |
| strictLC_popCtrl | Parkinsons | **0.08** | 0.22 | 0.36 | 0.72 |
| strictLC_infCtrl | Parkinsons | **-0.01** | 0.12 | -0.05 | 0.96 |
| C1 | Periph_neuro | **0.47** | 0.09 | 5.13 | 0 |
| C2 | Periph_neuro | **-0.03** | 0.09 | -0.37 | 0.71 |
| C3 | Periph_neuro | **0.29** | 0.09 | 3.09 | 0 |
| C4 | Periph_neuro | **0.59** | 0.09 | 6.37 | 0 |
| C5 | Periph_neuro | **-0.16** | 0.09 | -1.82 | 0.07 |
| C6 | Periph_neuro | **-0.52** | 0.07 | -7.19 | 0 |
| broadLC_popCtrl | Periph_neuro | **0.09** | 0.17 | 0.56 | 0.58 |
| broadLC_infCtrl | Periph_neuro | **0.16** | 0.19 | 0.86 | 0.39 |
| strictLC_popCtrl | Periph_neuro | **0.29** | 0.27 | 1.07 | 0.28 |
| strictLC_infCtrl | Periph_neuro | **0.20** | 0.11 | 1.76 | 0.08 |
| C1 | Prostate_carci | **-0.03** | 0.04 | -0.72 | 0.47 |
| C2 | Prostate_carci | **-0.05** | 0.05 | -0.95 | 0.34 |
| C3 | Prostate_carci | **0.06** | 0.05 | 1.14 | 0.25 |
| C4 | Prostate_carci | **-0.04** | 0.06 | -0.66 | 0.51 |
| C5 | Prostate_carci | **-0.07** | 0.06 | -1.30 | 0.19 |
| C6 | Prostate_carci | **0.08** | 0.04 | 1.97 | 0.05 |
| broadLC_popCtrl | Prostate_carci | **-0.21** | 0.17 | -1.27 | 0.21 |
| broadLC_infCtrl | Prostate_carci | **-0.25** | 0.17 | -1.51 | 0.13 |
| strictLC_popCtrl | Prostate_carci | **-0.22** | 0.24 | -0.92 | 0.36 |
| strictLC_infCtrl | Prostate_carci | **-0.09** | 0.07 | -1.36 | 0.17 |
| C1 | Scoliosis | **0.20** | 0.12 | 1.71 | 0.09 |
| C2 | Scoliosis | **0.17** | 0.13 | 1.24 | 0.22 |
| C3 | Scoliosis | **0.16** | 0.14 | 1.15 | 0.25 |
| C4 | Scoliosis | **0.23** | 0.14 | 1.73 | 0.08 |
| C5 | Scoliosis | **-0.26** | 0.12 | -2.20 | 0.03 |
| C6 | Scoliosis | **-0.21** | 0.10 | -2.06 | 0.04 |
| broadLC_popCtrl | Scoliosis | **0.24** | 0.31 | 0.77 | 0.44 |
| broadLC_infCtrl | Scoliosis | **0.19** | 0.30 | 0.63 | 0.53 |
| strictLC_popCtrl | Scoliosis | **0.33** | 0.49 | 0.68 | 0.50 |
| strictLC_infCtrl | Scoliosis | **0.16** | 0.17 | 0.89 | 0.37 |
| C1 | Systolic_HF | **0.51** | 0.07 | 7.27 | 0 |
| C2 | Systolic_HF | **0.81** | 0.10 | 8.31 | 0 |
| C3 | Systolic_HF | **0.19** | 0.08 | 2.36 | 0.02 |
| C4 | Systolic_HF | **-0.08** | 0.07 | -1.08 | 0.28 |
| C5 | Systolic_HF | **0.11** | 0.07 | 1.73 | 0.08 |
| C6 | Systolic_HF | **-0.67** | 0.06 | -11.11 | 0 |
| broadLC_popCtrl | Systolic_HF | **0.28** | 0.19 | 1.45 | 0.15 |
| broadLC_infCtrl | Systolic_HF | **0.20** | 0.18 | 1.06 | 0.29 |
| strictLC_popCtrl | Systolic_HF | **0.18** | 0.24 | 0.75 | 0.45 |
| strictLC_infCtrl | Systolic_HF | **-0.01** | 0.09 | -0.14 | 0.89 |

### Supplementary Table 5. Genome-wide genetic correlations between multimorbidity cluster memberships and two long COVID definitions

| **Trait1** | **Trait2** | **rg** | **se** | **z** | **p** |
| --- | --- | --- | --- | --- | --- |
| C1 | C1 | **1** | 0 | 1519800 | 0 |
| C1 | C2 | **0.51** | 0.12 | 4.10 | 0 |
| C1 | C3 | **0.47** | 0.15 | 3.14 | 0 |
| C1 | C4 | **0.18** | 0.13 | 1.40 | 0.16 |
| C1 | C5 | **-0.12** | 0.12 | -1.02 | 0.31 |
| C1 | C6 | **-0.85** | 0.06 | -13.34 | 0 |
| C1 | broadLC_infCtrl | **0.64** | 0.40 | 1.59 | 0.11 |
| C1 | broadLC_popCtrl | **0.71** | 0.47 | 1.51 | 0.13 |
| C1 | strictLC_infCtrl | **0.28** | 0.17 | 1.62 | 0.11 |
| C1 | strictLC_popCtrl | **0.92** | 0.77 | 1.18 | 0.24 |
| C2 | C1 | **0.51** | 0.12 | 4.10 | 0 |
| C2 | C2 | **1** | 0 | 25352.15 | 0 |
| C2 | C3 | **-0.06** | 0.15 | -0.43 | 0.67 |
| C2 | C4 | **-0.60** | 0.14 | -4.41 | 0 |
| C2 | C5 | **0.52** | 0.19 | 2.77 | 0.01 |
| C2 | C6 | **-0.61** | 0.08 | -7.60 | 0 |
| C2 | broadLC_infCtrl | **-0.02** | 0.30 | -0.07 | 0.94 |
| C2 | broadLC_popCtrl | **0.10** | 0.36 | 0.29 | 0.77 |
| C2 | strictLC_infCtrl | **0.14** | 0.18 | 0.78 | 0.44 |
| C2 | strictLC_popCtrl | **0.50** | 0.54 | 0.93 | 0.35 |
| C3 | C1 | **0.47** | 0.15 | 3.14 | 0 |
| C3 | C2 | **-0.06** | 0.15 | -0.43 | 0.67 |
| C3 | C3 | **1** | 0 | 46835.79 | 0 |
| C3 | C4 | **0.25** | 0.16 | 1.53 | 0.13 |
| C3 | C5 | **-0.26** | 0.14 | -1.89 | 0.06 |
| C3 | C6 | **-0.50** | 0.09 | -5.37 | 0 |
| C3 | broadLC_infCtrl | **0.28** | 0.36 | 0.77 | 0.44 |
| C3 | broadLC_popCtrl | **0.53** | 0.45 | 1.18 | 0.24 |
| C3 | strictLC_infCtrl | **0** | 0.19 | -0.01 | 0.99 |
| C3 | strictLC_popCtrl | **0.28** | 0.47 | 0.59 | 0.55 |
| C4 | C1 | **0.18** | 0.13 | 1.40 | 0.16 |
| C4 | C2 | **-0.60** | 0.14 | -4.41 | 0 |
| C4 | C3 | **0.25** | 0.16 | 1.53 | 0.13 |
| C4 | C4 | **1** | 0 | 587865.48 | 0 |
| C4 | C5 | **-0.69** | 0.12 | -5.80 | 0 |
| C4 | C6 | **-0.06** | 0.13 | -0.45 | 0.66 |
| C4 | broadLC_infCtrl | **0.59** | 0.41 | 1.43 | 0.15 |
| C4 | broadLC_popCtrl | **0.46** | 0.39 | 1.18 | 0.24 |
| C4 | strictLC_infCtrl | **0.22** | 0.19 | 1.18 | 0.24 |
| C4 | strictLC_popCtrl | **0.43** | 0.50 | 0.86 | 0.39 |
| C5 | C1 | **-0.12** | 0.12 | -1.02 | 0.31 |
| C5 | C2 | **0.52** | 0.19 | 2.77 | 0.01 |
| C5 | C3 | **-0.26** | 0.14 | -1.89 | 0.06 |
| C5 | C4 | **-0.69** | 0.12 | -5.80 | 0 |
| C5 | C5 | **1** | 0 | 44748.56 | 0 |
| C5 | C6 | **-0.27** | 0.10 | -2.80 | 0.01 |
| C5 | broadLC_infCtrl | **-0.53** | 0.39 | -1.37 | 0.17 |
| C5 | broadLC_popCtrl | **-0.54** | 0.40 | -1.35 | 0.18 |
| C5 | strictLC_infCtrl | **-0.16** | 0.18 | -0.93 | 0.35 |
| C5 | strictLC_popCtrl | **-0.39** | 0.47 | -0.83 | 0.41 |
| C6 | C1 | **-0.85** | 0.06 | -13.34 | 0 |
| C6 | C2 | **-0.61** | 0.08 | -7.60 | 0 |
| C6 | C3 | **-0.50** | 0.09 | -5.37 | 0 |
| C6 | C4 | **-0.06** | 0.13 | -0.45 | 0.66 |
| C6 | C5 | **-0.27** | 0.10 | -2.80 | 0.01 |
| C6 | C6 | **1** | 0 | 67435.09 | 0 |
| C6 | broadLC_infCtrl | **-0.39** | 0.32 | -1.23 | 0.22 |
| C6 | broadLC_popCtrl | **-0.49** | 0.38 | -1.28 | 0.20 |
| C6 | strictLC_infCtrl | **-0.22** | 0.13 | -1.59 | 0.11 |
| C6 | strictLC_popCtrl | **-0.75** | 0.64 | -1.18 | 0.24 |
| broadLC_infCtrl | C1 | **0.64** | 0.40 | 1.59 | 0.11 |
| broadLC_infCtrl | C2 | **-0.02** | 0.30 | -0.07 | 0.94 |
| broadLC_infCtrl | C3 | **0.28** | 0.36 | 0.77 | 0.44 |
| broadLC_infCtrl | C4 | **0.59** | 0.41 | 1.43 | 0.15 |
| broadLC_infCtrl | C5 | **-0.53** | 0.39 | -1.37 | 0.17 |
| broadLC_infCtrl | C6 | **-0.39** | 0.32 | -1.23 | 0.22 |
| broadLC_infCtrl | broadLC_infCtrl | **1** | 0 | 567483.27 | 0 |
| broadLC_infCtrl | broadLC_popCtrl | **1.85** | 0.83 | 2.23 | 0.03 |
| broadLC_infCtrl | strictLC_infCtrl | **1.94** | 0.64 | 3.05 | 0 |
| broadLC_infCtrl | strictLC_popCtrl | **4.32** | 3.53 | 1.22 | 0.22 |
| broadLC_popCtrl | C1 | **0.71** | 0.47 | 1.51 | 0.13 |
| broadLC_popCtrl | C2 | **0.10** | 0.36 | 0.29 | 0.77 |
| broadLC_popCtrl | C3 | **0.53** | 0.45 | 1.18 | 0.24 |
| broadLC_popCtrl | C4 | **0.46** | 0.39 | 1.18 | 0.24 |
| broadLC_popCtrl | C5 | **-0.54** | 0.40 | -1.35 | 0.18 |
| broadLC_popCtrl | C6 | **-0.49** | 0.38 | -1.28 | 0.20 |
| broadLC_popCtrl | broadLC_infCtrl | **1.85** | 0.83 | 2.23 | 0.03 |
| broadLC_popCtrl | broadLC_popCtrl | **1** | 0 | 8482.31 | 0 |
| broadLC_popCtrl | strictLC_infCtrl | **1.91** | 0.78 | 2.45 | 0.01 |
| broadLC_popCtrl | strictLC_popCtrl | **2.85** | 2.20 | 1.29 | 0.20 |
| strictLC_infCtrl | C1 | **0.28** | 0.17 | 1.62 | 0.11 |
| strictLC_infCtrl | C2 | **0.14** | 0.18 | 0.78 | 0.44 |
| strictLC_infCtrl | C3 | **0.00** | 0.19 | -0.01 | 0.99 |
| strictLC_infCtrl | C4 | **0.22** | 0.19 | 1.18 | 0.24 |
| strictLC_infCtrl | C5 | **-0.16** | 0.18 | -0.93 | 0.35 |
| strictLC_infCtrl | C6 | **-0.22** | 0.13 | -1.59 | 0.11 |
| strictLC_infCtrl | broadLC_infCtrl | **1.94** | 0.64 | 3.05 | 0 |
| strictLC_infCtrl | broadLC_popCtrl | **1.91** | 0.78 | 2.45 | 0.01 |
| strictLC_infCtrl | strictLC_infCtrl | **1** | 0 | 9393.95 | 0 |
| strictLC_infCtrl | strictLC_popCtrl | **2.24** | 1.48 | 1.51 | 0.13 |
| strictLC_popCtrl | C1 | **0.92** | 0.77 | 1.18 | 0.24 |
| strictLC_popCtrl | C2 | **0.50** | 0.54 | 0.93 | 0.35 |
| strictLC_popCtrl | C3 | **0.28** | 0.47 | 0.59 | 0.55 |
| strictLC_popCtrl | C4 | **0.43** | 0.50 | 0.86 | 0.39 |
| strictLC_popCtrl | C5 | **-0.39** | 0.47 | -0.83 | 0.41 |
| strictLC_popCtrl | C6 | **-0.75** | 0.64 | -1.18 | 0.24 |
| strictLC_popCtrl | broadLC_infCtrl | **4.32** | 3.53 | 1.22 | 0.22 |
| strictLC_popCtrl | broadLC_popCtrl | **2.85** | 2.20 | 1.29 | 0.20 |
| strictLC_popCtrl | strictLC_infCtrl | **2.24** | 1.48 | 1.51 | 0.13 |
| strictLC_popCtrl | strictLC_popCtrl | **1** | 0 | 4422.76 | 0 |

### Supplementary Table 6. Results of local genetic correlation analysis between long COVID phenotypes and the multimorbidity clusters.

#### A. Bivariate local genetic correlation summary between strict long COVID/infected control and multimorbidity clusters

| **pair** | **n_bivar** | **n_nominal_0.05** | **n_FDR_pair_0.05** | **min_p** | **top_chr** | **top_start** | **top_stop** | **top_rho_ci** |
| --- | --- | --- | --- | --- | --- | --- | --- | --- |
| **strictLC_infCtrl**  **~ C1** | 3 | 0 | 0 | 0.17 | 16 | 88387229 | 89106304 | -0.506  (-1.000, 0.270) |
| **strictLC_infCtrl**  **~ C2** | 4 | 0 | 0 | 0.13 | 16 | 88387229 | 89106304 | -0.653  (-1.000, 0.214) |
| **strictLC_infCtrl**  **~ C4** | 3 | 0 | 0 | 0.43 | 1 | 1376205 | 2215495 | 0.253  (-0.475, 1.000) |
| **strictLC_infCtrl**  **~ C5** | 10 | 0 | 0 | 0.14 | 4 | 96183802 | 97540787 | 0.489  (-0.200, 1.000) |

#### B. Bivariate local genetic correlation summary between broad long COVID/infected control and multimorbidity clusters

| **pair** | **n_bivar** | **n_nominal_0.05** | **n_FDR_pair_0.05** | **min_p** | **top_chr** | **top_start** | **top_stop** | **top_rho_ci** |
| --- | --- | --- | --- | --- | --- | --- | --- | --- |
| **broadLC_infCtrl ~ C1** | 16 | 0 | 0 | 0.07 | 16 | 88387229 | 89106304 | -0.500  (-1.000, 0.045) |
| **broadLC_infCtrl ~ C2** | 21 | 2 | 0 | 0.01 | 15 | 90632719 | 91593623 | -1.000  (-1.000,  -0.151) |
| **broadLC_infCtrl ~ C3** | 18 | 0 | 0 | 0.10 | 7 | 71215742 | 73128830 | -0.582  (-1.000, 0.139) |
| **broadLC_infCtrl ~ C4** | 16 | 1 | 0 | 0.01 | 3 | 85807680 | 86660979 | 1.000 (0.202, 1.000) |
| **broadLC_infCtrl ~ C5** | 21 | 1 | 0 | 0.02 | 9 | 105846546 | 107061019 | -0.949  (-1.000,  -0.151) |
| **broadLC_infCtrl ~ C6** | 29 | 1 | 0 | 0.04 | 2 | 3314203 | 4016104 | 0.547 (0.043, 1.000) |

#### C. Bivariate local genetic correlation summary between strict long COVID/population control and multimorbidity clusters

| **pair** | **n_bivar** | **n_nominal_0.05** | **n_FDR_pair_0.05** | **min_p** | **top_chr** | **top_start** | **top_stop** | **top_rho_ci** |
| --- | --- | --- | --- | --- | --- | --- | --- | --- |
| **C1 ~ strictLC_popCtrl** | 42 | 4 | 0 | 0.01 | 11 | 45019560 | 46316005 | -0.822  (-1.000,  -0.241) |
| **C2 ~ strictLC_popCtrl** | 44 | 2 | 0 | 0.02 | 19 | 54602371 | 55182973 | 0.849  (0.176, 1.000) |
| **C3 ~ strictLC_popCtrl** | 32 | 1 | 0 | 0.00 | 8 | 38803981 | 40179577 | 0.949  (0.320, 1.000) |
| **C4 ~ strictLC_popCtrl** | 38 | 3 | 0 | 0.00 | 16 | 89106305 | 90292787 | 0.908  (0.311, 1.000) |
| **C5 ~ strictLC_popCtrl** | 42 | 1 | 0 | 0.05 | 14 | 56206431 | 57460781 | 0.654  (0.017, 1.000) |
| **C6 ~ strictLC_popCtrl** | 56 | 3 | 0 | 0.01 | 20 | 24487129 | 25181038 | -1.000  (-1.000,  -0.258) |

#### D. Bivariate local genetic correlation summary between broad long COVID/population control and multimorbidity clusters

| **pair** | **n_bivar** | **n_nominal_0.05** | **n_FDR_pair_0.05** | **min_p** | **top_chr** | **top_start** | **top_stop** | **top_rho_ci** |
| --- | --- | --- | --- | --- | --- | --- | --- | --- |
| **broadLC_popCtrl ~ C1** | 39 | 0 | 0 | 0.07 | 12 | 8123318 | 8760628 | -0.763  (-1.000, 0.066) |
| **broadLC_popCtrl ~ C2** | 39 | 3 | 0 | 0.01 | 1 | 163854568 | 164726579 | -0.998  (-1.000,  -0.290) |
| **broadLC_popCtrl ~ C3** | 29 | 2 | 0 | 0.01 | 4 | 61971162 | 63337954 | 0.962 (0.259, 1.000) |
| **broadLC_popCtrl ~ C4** | 35 | 1 | 0 | 0.01 | 9 | 33194893 | 34704544 | 1.000 (0.283, 1.000) |
| **broadLC_popCtrl ~ C5** | 41 | 2 | 0 | 0.03 | 20 | 24487129 | 25181038 | -0.783  (-1.000,  -0.102) |
| **broadLC_popCtrl ~ C6** | 46 | 3 | 0 | 0.00 | 10 | 10081280 | 10969481 | -1.000  (-1.000,  -0.388) |

### Supplementary Note: Extended Methods and Results

**Methods**

*S1. UK Biobank data fields*

Participant disease data was extracted from Hospital Episode Statistics using the following fields:

- Hospital inpatient: category 2000, subcategory 2002, fields 41204, 41270 and 41280
- Death register: category 100093, fields 40001 and 40002
- Cancer register: category 100092, field 40006

*S2. Local genetic correlation analysis (LAVA)*

Pairwise phenotypic correlations were computed and compared with genome-wide genetic correlations estimated with LDSC. Local genetic correlations were estimated using the **L**ocal **A**nalysis of [co]**V**ariant **A**ssociation (LAVA) package^1^, applied in an agnostic approach covering the whole genome systematically. LAVA first performed univariate tests to detect loci showing significant local heritability for each phenotype separately, then estimated local genetic correlations between pairs of traits within loci significant for both via bivariate tests. Only loci showing consistent, correlated SNP effects across both traits were retained. Pre-computed European ancestry LD blocks (minimum block size 2,500 kb, GRCh37 build) and UK Biobank LD reference panels (100,000 unrelated UK Biobank European participants, LD calculated up to 10 Mb per SNP) were used. The analysis was adjusted for sample overlap derived from global genetic correlation intercepts ^1,2^. Regions were gene-mapped using Ensembl BioMart and functionally annotated ^3^.

*S3. Colocalisation prior probabilities*

Default coloc prior probabilities were used (p_1_ = 10^-4^, p_2_ = 10^-4^, p_12_ = 10^-5^).

*S4. Finemapping parameters*

Fine-mapping analyses were performed using FINEMAP (v1.4) to refine the shared causal signal identified via colocalisation and prioritise variants most likely to drive the association within this region^4^. Analyses were conducted separately for each relevant cluster within a ±1 Mb window centred on the lead variant we investigated. FINEMAP was run allowing for up to five causal variants per region (default configuration). SNP-level summary statistics were provided as z-score files, and linkage disequilibrium (LD) matrices were computed from the same UK Biobank reference sample using PLINK (r values, square matrix). Default FINEMAP settings were used, including a correlation threshold of 0.95 for grouping configurations and convergence assessed over 100 iterations with a posterior probability tolerance of 0.001. Evidence for candidate causal variants was assessed using posterior inclusion probabilities (PIPs) and credible sets.

*S5. Gene-based association analysis (MAGMA)*

Gene-based association analyses were performed using MAGMA, which aggregates SNP-level associations into gene-level statistics while accounting for linkage disequilibrium^5^. Tests were run separately for each multimorbidity cluster using GWAS summary statistics, with SNPs mapped to genes based on the NCBI37.3 gene location reference (GRCh37 coordinates) and a UK Biobank LD reference panel derived from biallelic autosomal variants. Default MAGMA parameters were used. Gene-level significance was assessed using false discovery rate correction (FDR < 0.05).

*S6. Long COVID summary statistics*

The long COVID GWAS summary statistics comprised four case-control definitions with the following sample sizes: strict case vs population control (3,018 cases, 994,582 controls; total N = 997,600), broad case vs population control (6,450 cases, 1,093,995 controls; total N = 1,100,445), strict case vs strict control (2,964 cases, 37,935 COVID-19-positive controls; total N = 40,899), and broad case vs strict control (6,396 cases, 46,208 COVID-19-positive controls; total N = 52,604). Strict cases were individuals with test-verified SARS-CoV-2 infection; broad cases did not require test confirmation of acute infection. Strict controls had test-confirmed SARS-CoV-2 infection but did not develop long COVID; broad controls were population controls who did not meet long COVID criteria. Although the meta-analyses were multi-ancestry, they were predominantly composed of individuals of European ancestry; therefore, European ancestry LD reference panels were used in all downstream analyses.

**Results**

*S7. Cluster demographic characteristics*

Sex distribution differed across clusters, with higher proportions of females observed in the Gastrointestinal (C3), Musculoskeletal (C4), and Cancers & Eye Disorders (C6) clusters. Mean age at assessment was similar across clusters, ranging from 67.5 to 68.4 years. Marked differences were observed in disease burden and mortality: clusters characterised by neurodegenerative and cardiovascular conditions showed higher mean numbers of disease categories and higher mortality rates, whereas clusters with lower overall disease burden, such as Musculoskeletal (C4) and Cancers & Eye Disorders (C6), exhibited comparatively lower mortality. The Vascular cluster (C5) displayed a relatively elevated mortality rate despite a lower mean number of disease categories, suggesting heterogeneity in risk profiles across clusters.
