## Extended Data for "Exploring the genetic architecture of multimorbidity and its impact on long COVID risk"

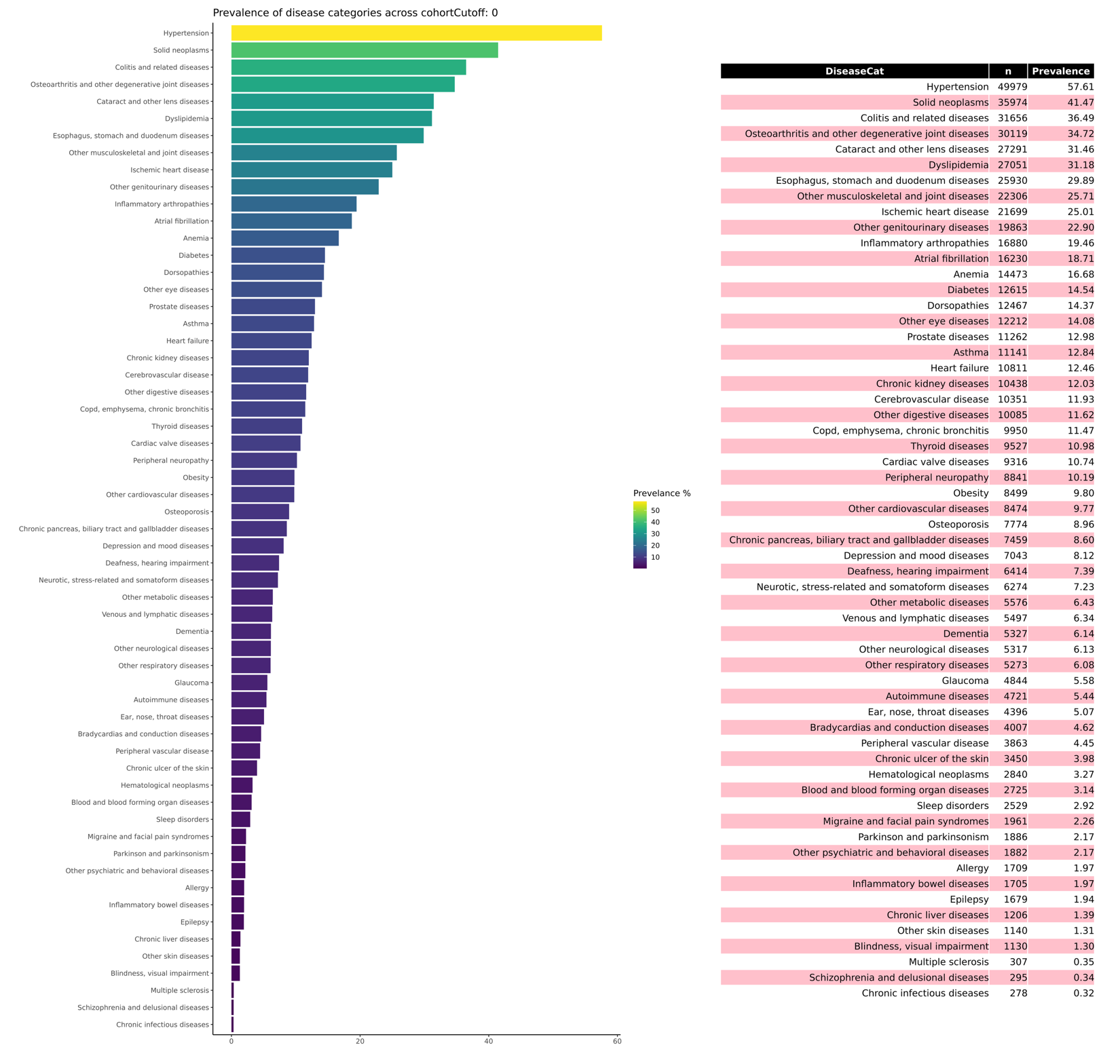

### Extended Data Figure 1: Prevalence of 59 disease categories in the multimorbid cohort

Extended Data Figure 2: Average clustering quality index scores for fuzzy c-means clustering parameter optimisation. Indices were evaluated across K = 2- 12 clusters and fuzzifier values m = 1.1-1.5 and averaged over 100 realisations. (A) Calinski-Harabasz index, (B) Partition Entropy, (C) Partition Coefficient, and (D) Fukuyama-Sugeno index. The steepest change in clustering quality was observed between K = 4 to K = 7, and the final choice of K was further guided by the interpretability of cluster composition.

(A)
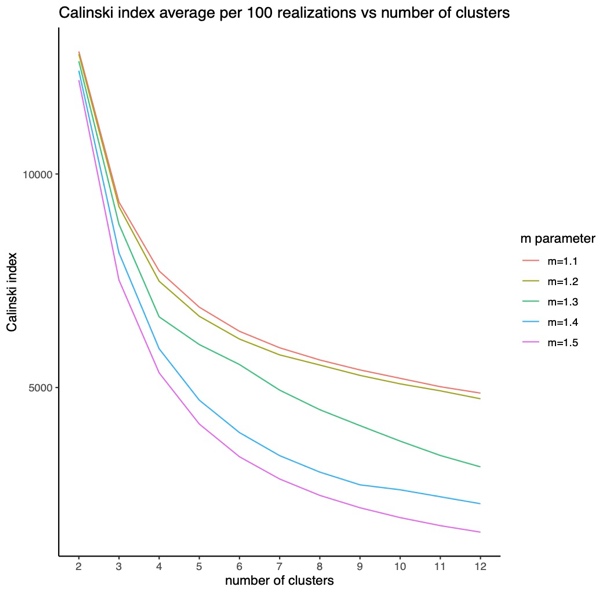
(B)
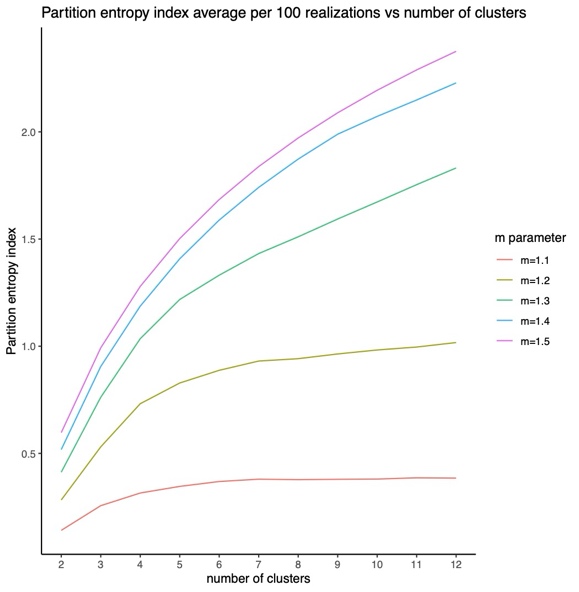

(C)
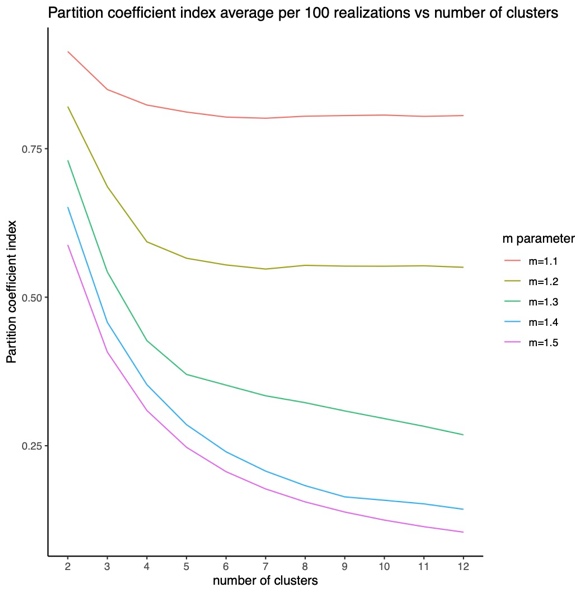
(D)
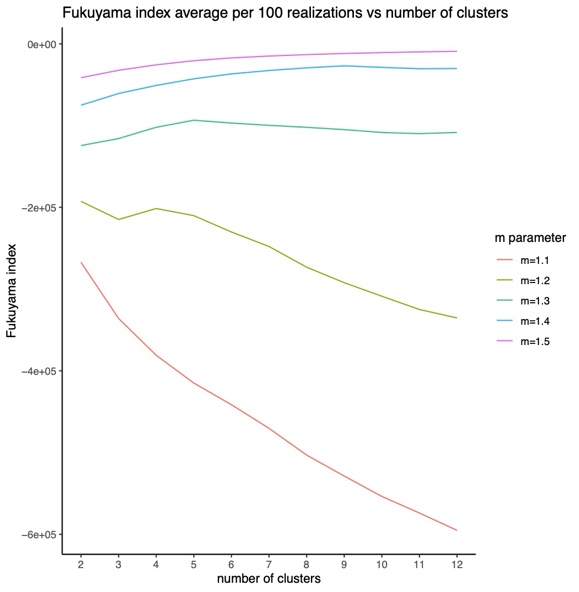

Note: After evaluating clustering quality across different combinations of cluster numbers and fuzzifier values, we selected *K* = 6 clusters at *m* = 1.2. Disease categories were assigned to clusters based on exclusivity and observed-to-expected prevalence ratios within each cluster. Clusters were named according to their dominant disease pattern: Neurodegenerative (C1), Cardiac (C2), Gastrointestinal (C3), Musculoskeletal (C4), Vascular (C5), and Cancers & Eye disorders (C6).

Extended Data Figure 3: Genetic coherence heatmap of MM clusters vs hallmark diseases. Cells show genetic correlation (rg) (colour scale centred at 0). Asterisks mark FDR < 0.05 (Benjamini–Hochberg) across all tests within the panel. IHD: Ischemic Heart Disease, Systolic hf: Systolic heart failure, IBD: Inflammatory Bowel Disease, Prostate carci: prostate carcinoma, Periph neuro: peripheral neuropathy.

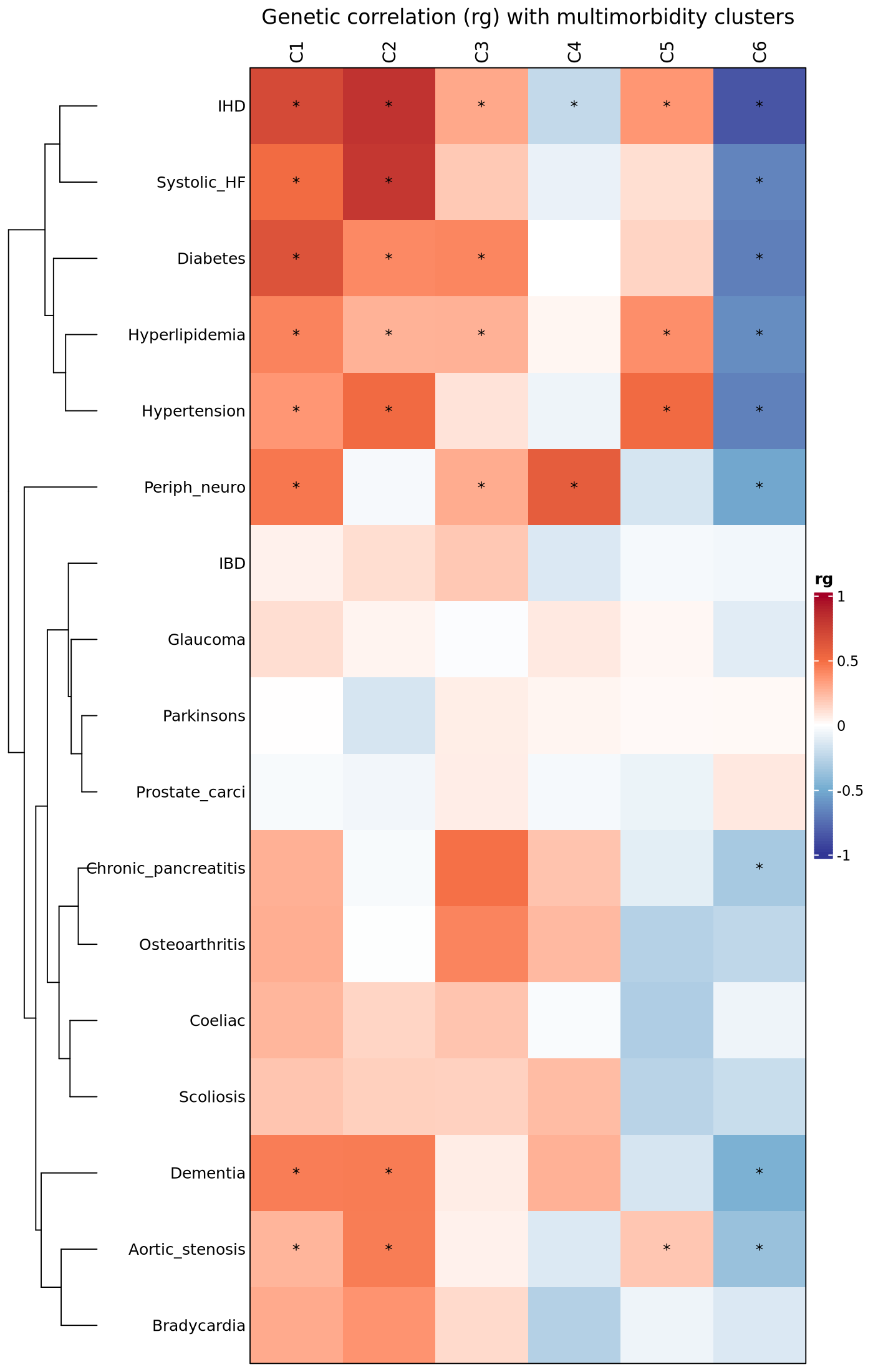

Note: Clusters demonstrated the expected pattern of positive genetic correlations with their hallmark diseases, including strong correlations between the Neurodegenerative cluster (C1) and dementia or peripheral neuropathies, the Cardiac cluster (C2) and ischaemic heart disease, hypertension and hyperlipidaemia, and between the Vascular cluster (C5) and diabetes and systolic hypertension (FDR < 0.05). Correlations with non-hallmark conditions were attenuated, supporting phenotype validity and specificity. Notably, the Cancers & Eye disorders cluster (C6) showed broadly inverse genetic correlations across multiple clusters and hallmark diseases, suggesting a distinct and opposing genetic architecture relative to cardiometabolic and neurodegenerative multimorbidity.

Extended Data Figure 4: Genetic coherence heatmap of long COVID vs hallmark diseases. Cells show genetic correlation (rg) (colour scale centred at 0). Asterisks mark FDR < 0.05 (Benjamini–Hochberg) across all tests within the panel. IHD: Ischemic Heart Disease, Systolic HF: Systolic heart failure, IBD: Inflammatory Bowel Disease, Prostate carci: prostate carcinoma, Periph neuro: peripheral neuropathy.

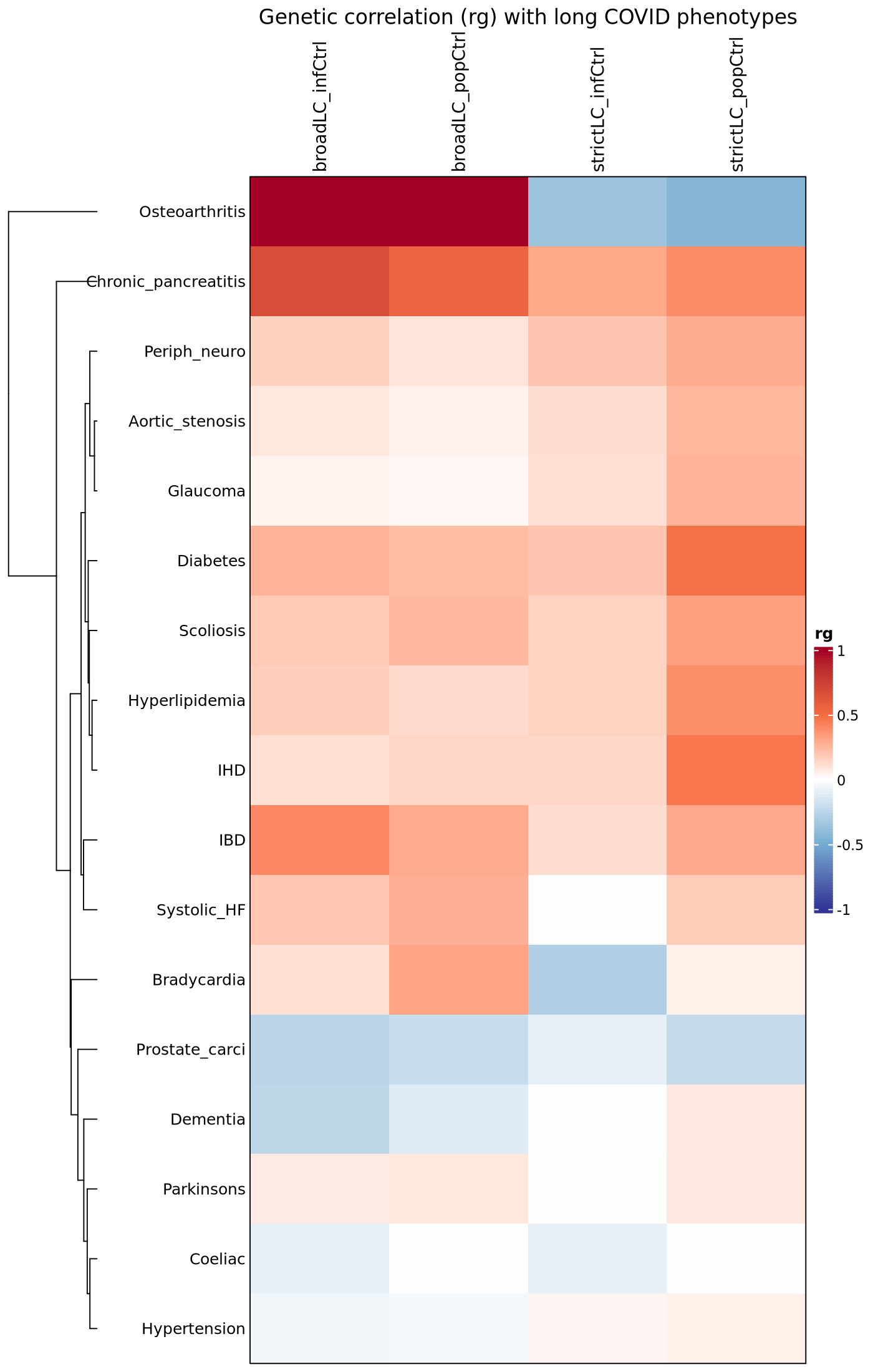

Note: Genome-wide genetic correlations between strict and broad long COVID case definitions and hallmark diseases were computed using LDSC (Figure 7; Supplemental Table 6). None were statistically significant. While correlation profiles were mostly concordant across the two definitions, several disease categories diverged markedly. For example, osteoporosis or bradycardia displayed opposite directions and discordant effect sizes between the strict and broad long COVID definitions. These discrepancies highlight that genetic correlation estimates vary depending on the long COVID case definition used. Given the definitional variability of long COVID, we treat these findings as exploratory only.

Extended Data Table 1. Multimorbidity cluster membership-associated variants identified in the conditional and joint association analysis. Table headers are: Chr: chromosome; SNP: SNP rsID; bp: physical position; refA: effect allele, freq: frequency of the effect allele in the original data; b: effect size, se: standard error, p: p-value from the original GWAS; freq_geno: frequency of the effect allele in the reference sample; bJ: effect size of the joint analysis of all the selected SNPs, bJ_se: corresponding standard error, pJ: p-value from the joint analysis of all the selected SNPs; LD_r: LD correlation between the SNP i and SNP i + 1 for the SNPs on the list.

| **Trait** | **Chr** | **SNP** | **bp** | **refA** | **freq** | **b** | **se** | **p** | **freq_geno** | **bJ** | **bJ_se** | **pJ** | **LD_r** |
| --- | --- | --- | --- | --- | --- | --- | --- | --- | --- | --- | --- | --- | --- |
| **C1** | 19 | rs429358 | 45411941 | T | 0.85 | -0.006 | 0.001 | 2.6E-09 | 0.85 | -0.006 | 0.001 | 2.6E-09 | 0 |
| **C2** | 4 | Rs72900157 | 111710793 | C | 0.90 | -0.011 | 0.002 | 1.5E-10 | 0.9 | -0.011 | 0.002 | 1.5E-10 | 0 |
| **C2** | 6 | Rs55730499 | 161005610 | C | 0.92 | -0.015 | 0.002 | 1.1E-15 | 0.92 | -0.015 | 0.002 | 1.2E-15 | 0 |
| **C2** | 9 | rs10757277 | 22124450 | A | 0.52 | -0.006 | 0.001 | 1.2E-08 | 0.52 | -0.006 | 0.001 | 1.2E-08 | 0 |
| **C4** | 19 | Rs1065853 | 45413233 | G | 0.92 | -0.012 | 0.002 | 2.4E-08 | 0.92 | -0.012 | 0.002 | 2.4E-08 | 0 |
| **C5** | 19 | Rs1065853 | 45413233 | G | 0.92 | 0.016 | 0.002 | 7.4E-14 | 0.92 | 0.016 | 0.002 | 7.5E-14 | 0 |
| **C6** | 4 | Rs71611783 | 175315167 | A | 0.98 | -0.032 | 0.005 | 3.3E-09 | 0.98 | -0.032 | 0.005 | 3.3E-09 | 0 |

### Extended Data Table 2: FDR-significant LAVA loci shared amongst MM clusters.

| **Cluster Pair** | **chr** | **start** | **stop** | **rho** | **rho.lower** | **rho.upper** | **p** | **p_FDR_pair** |
| --- | --- | --- | --- | --- | --- | --- | --- | --- |
| **C1-C5** | 6 | 91097025 | 92015279 | -0.85 | -1 | -0.16 | 0.033 | 0.033 |
| **C3-C6** | 2 | 65938003 | 67224027 | -1 | -1 | -0.41 | 0.007 | 0.029 |
| **C3-C6** | 6 | 32897999 | 33194975 | -0.84 | -1 | -0.22 | 0.036 | 0.049 |
| **C3-C6** | 6 | 66445496 | 67036787 | -0.96 | -1 | -0.36 | 0.026 | 0.049 |
| **C4-C6** | 3 | 68319771 | 69784969 | -0.95 | -1 | -0.49 | 0.004 | 0.014 |

Note: Local genetic correlation using LAVA identified 13 genomic regions showing nominal significance, of which 5 remained significant after FDR correction (FDR < 0.05). A strong inverse local genetic correlation was observed between C4 and C6 on chromosome 3 (ρ = −0.95, 95% CI [−1.00, −0.49], P = 3.5 × 10⁻³). This region harbours TAFA1 (FAM19A1), TAFA4 (FAM19A4) and EOGT, implicating immune regulation and proliferative signalling pathways in the inverse comorbidity mechanism between these clusters^55–59^. Additional significant negative correlations were detected between C3 and C6 in loci on chromosomes 2 and 6. The chromosome 2 locus (ρ = –1.00, 95% CI [-1.00, -0.41], P = 7.4 x 10⁻³) mapped to MEIS1, a developmental transcription factor involved in haematopoiesis and leukemogenesis^60,61^. The locus on chromosome 6 spanned the major histocompatibility complex class II region and contained multiple immune-related genes, including HLA-DMA, HLA-DMB, HLA-DPA1, HLA-DPB1, and HLA-DOA, as well as BRD2 and RING1. In contrast, a second chromosome 6 region contained only pseudogenes (e.g. SLC25A51P1, ADH5P4, NUFIP1P). A signal between C1 and C5 clusters was observed on chromosome 6 (ρ = -0.85, 95% CI [-1.00, -0.16], P = 0.033) and mapped to MAP3K7, encoding TAK1, a kinase involved in inflammatory and stress-response pathways^62,63^. These results suggest that inverse correlations between multimorbidity clusters may be underpinned by specific genomic regions where allele effects act in opposite directions.

Extended Data Table 3. Finemapping of chromosome 19 locus shared by C4 and C5. SNPs highlighted in grey are shared amongst the two phenotypes. Multiple credible sets reflect uncertainty due to extreme linkage disequilibrium within the locus and do not indicate independent causal signals.

A. Results for C4 phenotype

| **Credible set** | **SNP** | **PIP** |
| --- | --- | --- |
| CS1 | rs4803743 | 1.00 |
| CS2 | rs1551891 | 0.76 |
| CS2 | rs62117161 | 0.24 |
| CS3 | rs2927487 | 1.00 |
| CS4 | rs11878852 | 1.00 |
| CS5 | rs181504313 | 0.98 |

B. Results for C5 phenotype

| **Credible set** | **SNP** | **PIP** |
| --- | --- | --- |
| CS1 | rs4803743 | 1.00 |
| CS2 | rs1551891 | 0.62 |
| CS2 | rs62117162 | 0.31 |
| CS2 | rs62117161 | 0.08 |
| CS3 | rs11878852 | 1.00 |
| CS4 | rs2927487 | 1.00 |
| CS5 | rs73037426 | 1.00 |

Note: Most shared variants showed opposite directions of effect on cluster membership, including rs4803743 (β = 0.0032 in cluster 4; β = −0.0044 in cluster 5), rs1551891, rs62117161 and rs11878852, consistent with the inverse genetic correlation observed between cluster 4 and 5 memberships. In contrast, one shared variant (rs2927487) showed concordant effect directions but was not significantly associated with either cluster. These findings indicate that the shared locus is characterised by antagonistic SNP-level effects across clusters, with inverse associations driven by variants acting in opposing directions rather than by a single concordant effect. No protein-coding variants were prioritised, indicating that the shared signal is likely mediated by regulatory mechanisms within the broader APOE–APOC1 haplotype block rather than by direct effects on protein structure.
